# From subthalamic local field potentials to the selection of chronic deep brain stimulation contacts in Parkinson’s disease - A systematic review

**DOI:** 10.1101/2024.10.29.24316351

**Authors:** Marjolein Muller, Mark F.C. van Leeuwen, Carel F. Hoffmann, Niels A. van der Gaag, Rodi Zutt, Saskia van der Gaag, Alfred C. Schouten, M. Fiorella Contarino

**Author notes:** Corresponding author, **Contact information corresponding author:** <u>E-mail</u>; <u>Postal Adress</u> dr. M.F.Contarino, Department of Neurology, Leiden University Medical Centre, Albinusdreef 2, 2333 ZA Leiden, the Netherlands. **Declarations of interest:** MFC is an independent consultant for research and educational issues of Medtronic (all fees to institution), is an independent consultant for research by INBRAIN (all fees to institution), provides research support/contracted research for Boston Scientific (all fees to institution) and received speaking fees for: ECMT (CME activity), and Boston Scientific (all fees to institution). All other authors declare that they have no known competing financial interests or personal relationships that could have appeared to influence the work reported in this paper.

## Abstract

**Background:** Programming deep brain stimulation (DBS) of the subthalamic nucleus for optimal symptom control in Parkinson’s Disease (PD) requires time and trained personnel. Novel implantable neurostimulators allow local field potentials (LFP) recording, which could be used to identify the optimal (chronic) stimulation contact. However, literature is inconclusive on which LFP features and prediction techniques are most effective.

**Objective:** To evaluate the performance of different LFP-based physiomarkers for predicting the optimal (chronic) stimulation contacts.

**Methods:** A literature search was conducted across nine databases, resulting in 418 individual papers. Two independent reviewers screened the articles based on title, abstract, and full text. The quality of included studies was assessed using a modified Joanna Briggs Institute Critical Appraisal Checklist for Case Series. Results were categorised in four classes based on the predictive performance with respect to the *a priori* chance.

**Results:** Twenty-five studies were included. Single-feature beta-band predictions demonstrated positive performance scores in 94% of the outcomes. Predictions based on single non-beta-frequency features yielded positive scores in only 25% of the outcomes, with positive results mainly for high frequency oscillations. Multi-feature predictions (e.g. machine learning) achieved accuracy scores within the two highest performance classes more often than single beta-based predictions (100% versus 39%).

**Conclusion:** Predicting the optimal stimulation contact based on LFP recordings is feasible and can improve DBS programming efficiency in PD. Single beta-band predictions show more promising results than non-beta-frequency features alone, but are outperformed by multi-feature predictions. Future research should further explore multi-feature predictions for optimal contact identification.

## Introduction

Deep brain stimulation (DBS) of the subthalamic nucleus (STN) is an effective treatment for patients with Parkinson’s disease (PD)[1, 2].

An important determinant of DBS effect, next to accurate implantation, is appropriate DBS programming, which starts by selection of the stimulation contact(s). Currently, this contact selection is often achieved by performing a monopolar review (MPR) [3]. During MPR, the thresholds for stimulation effects, including symptom reduction and side effects, are individually assessed for each contact (or ring in case of segmented levels) by clinical evaluation, while gradually increasing the stimulation amplitude. Contacts are then ranked and chosen for chronic stimulation based on their clinical efficacy and therapeutic window. This contact selection process is time-consuming (20-40 minutes per hemisphere), requires highly trained personnel, and can be exhausting and uncomfortable for patients [4, 5]. This is especially true for the currently available directional leads which contain eight individual contacts, and for other existing and future lead designs which may include even more contacts [6–8].

The results of a MPR can be influenced by several confounding factors such as patient fatigue, patient feedback, and the fact that some therapeutic effects of DBS have a latency period. [9]. In addition, the ‘stun’ effect resulting from lead implantation can temporarily decrease or even resolve the patient’s motor symptoms, complicating the MPR process [9]. Subsequently, the MPR is further complemented by an iterative process of stimulation and medication adjustments on an outpatient basis, which can take up to twelve months [10]. Hence, a faster and more objective solution is required.

One potentially promising technique is using the local field potential (LFP) recorded from the target structure [1, 11]. LFPs are electrical signals generated by the summed and synchronous electrical activity of individual neurons. The recorded signals are often interpreted in the frequency domain and can be recorded in a monopolar or bipolar manner using one or two electrodes on each implanted lead. Novel implantable neurostimulators, introduced in 2020, even allow for chronic LFP recording, simultaneously to stimulation. (figure 1).

**Figure 1.**
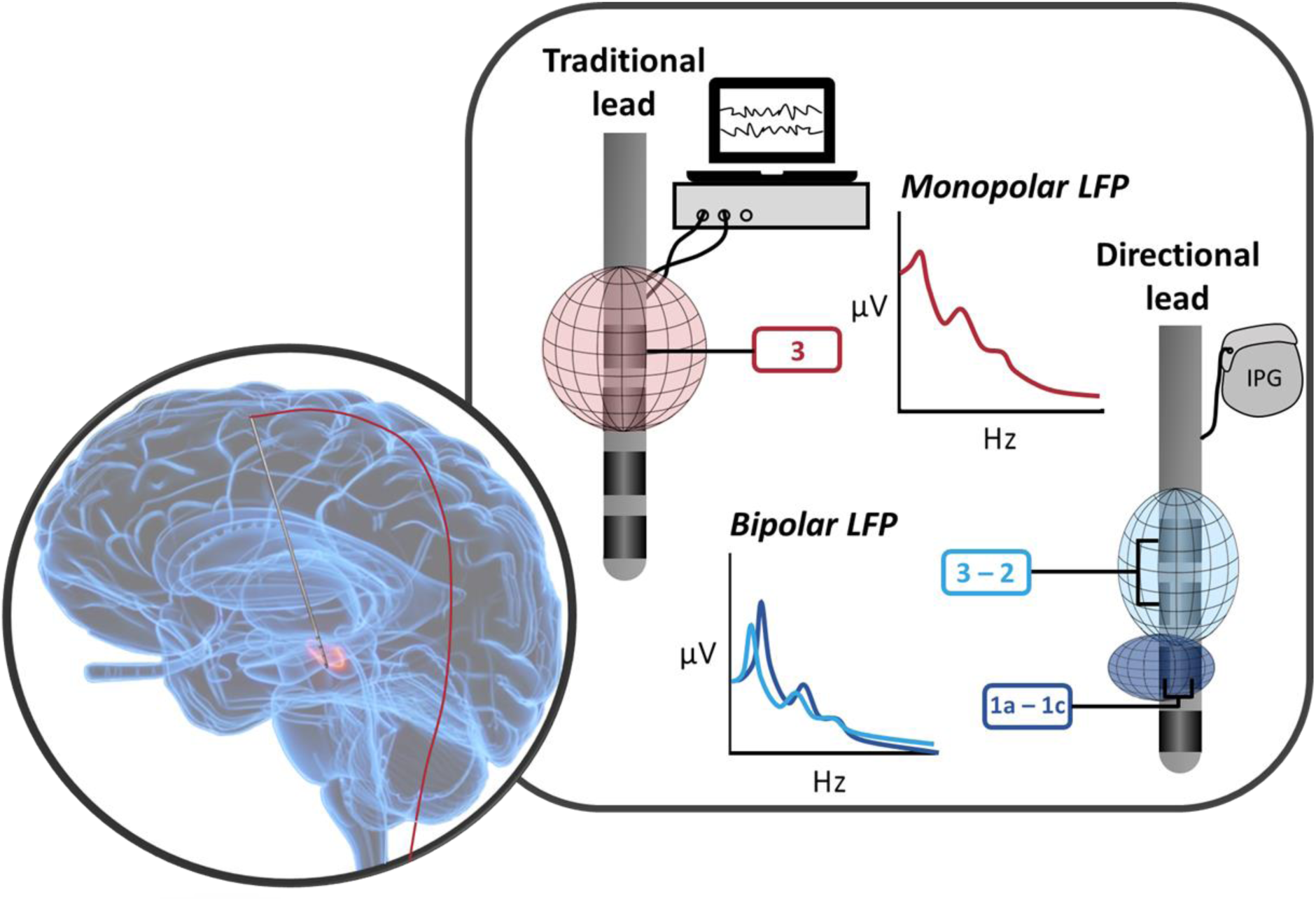
Schematic representation of possible techniques for local field potential (LFP) recording. LFPs can be recorded from the implanted traditional (4 cylindrical ‘ring’ electrodes) or directional lead (2 cylindrical ‘ring’ electrodes surrounding 2 levels split into 3 seperate ‘segment’ electrodes). Monopolar recordings can be obtained by means of an external system connected with externalised extensions; bipolar recordings can also be obtained by means of an implantable pulse generator (IPG).

For LFP recordings performed within the STN, one frequency band in particular, the beta-frequency band (13-35Hz), has shown a strong correlation with PD motor symptoms [12, 13]. Several studies also indicated that delivering stimulation through contacts closest to the beta source is often clinically most effective [12, 14], and that suppression of beta-activity is associated with improvement of motor symptoms in PD [12]. It is therefore hypothesised that beta LFPs could assist and optimise the DBS contact selection process [1, 11]. More recently, it has been suggested that other frequency bands may also be useful in the identification of the best contact for stimulation and that a combination of information from different frequency bands (e.g. theta (4-7Hz), alpha (8-12Hz), low-/high-beta (13-20/21-35Hz), gamma (40-100Hz), high frequency oscillations (HFO, 200-400Hz)) or non-frequency sources, such as anatomical landmarks or evoked resonant neural activity (ERNA) might increase the efficiency [15–17].

The objective of this systematic review was to evaluate the reported performance of different LFP-based neurophysiological biomarkers (physiomarkers) for predicting the optimal (chronic) stimulation contacts.

## Methods

### Search strategy

The literature search was conducted in accordance with the Preferred Reporting Items for Systematic Reviews and Meta-Analysis (PRISMA) 2009 guideline [18]. The search was updated until November 1^st^ , 2023. The following databases were included in our search: Pubmed, Embase, Web of Science, Cochrane Library, Emcare, PsycINFO, Academic Search Premier and Google Scholar. In Google Scholar only the first 50 most relevant articles were considered. All search terms covered three main subjects: PD; LFPs/neuronal oscillations and DBS programming. The complete search strings are reported in the supplementary methods (Supplementary file 1).

### Study selection

Two independent reviewers (MM and MvL) screened the articles. Any discrepancies were resolved through discussion. The following in- and exclusion criteria were used for the selection:

- <u>Inclusion criteria</u>: studies involving patients with PD; reporting of clinical choice or effect of stimulation contact; and seperately reporting LFP recordings from multiple contacts/contact pairs from leads implanted in the STN.
- <u>Exclusion criteria</u>: non-English articles; non-human evaluations (animal or computer simulations); literature review articles; case-reports; and non-peer reviewed manuscripts.

### Quality assessment

Two reviewers (MM and MvL) independently evaluated the methodological quality of each selected study using a self-adapted 17-points version of the Joanna Briggs Institute (JBI) Critical Appraisal Checklist for Case Series (Supplementary file 2) [19], in which questions regarding blinding, patient medication/activity state, and LFP acquisition, pre-processing and analysis techniques were added to better suit the purpose of this review.

### Data analysis

The reported LFP-based performance in predicting the optimal (chronic) stimulation contact, as identified through clinical evaluation (*clinical stimulation contact*), was transformed into a custom performance score to facilitate better comparisons across studies. First, information considering the predicted contact, the *clinical stimulation contact,* and the predictive performance was extracted from the included articles (figure 2, step 1). Second, the *a priori* chance of choosing the correct *optimal stimulation contact* was determined (figure 2, step 2). The *a priori* chance is based on the number of contact points on the lead and the amount of predicted contacts; for example, the *a priori* chance differs between cases with a directional lead containing 8 contacts (2 cylindrical ‘ring’ electrodes surrounding 2 levels split into 3 seperate ‘segment’ electrodes) and a traditional lead containing 4 contacts (4 ‘cylindrical’ ring electrodes). Third, the *multiplication factor* between the *a priori* chance and the predictive performance was determined (figure 2, step 3). Finally, the performance score could be assigned, allowing the following options: High performance: *factor* >3 times *a priori* chance; Moderate performance: *factor* ≤3 times but >2 times *a priori* chance OR significant correlation with p<0.005; Low performance: *factor* ≤2 times but > 1 time *a priori* chance OR significant correlation with p<0.05; Poor performance: *factor* ≤ 1 time *a priori* chance OR non-significant correlation with p>0.05 (figure 2, step 4).

**Figure 2.**
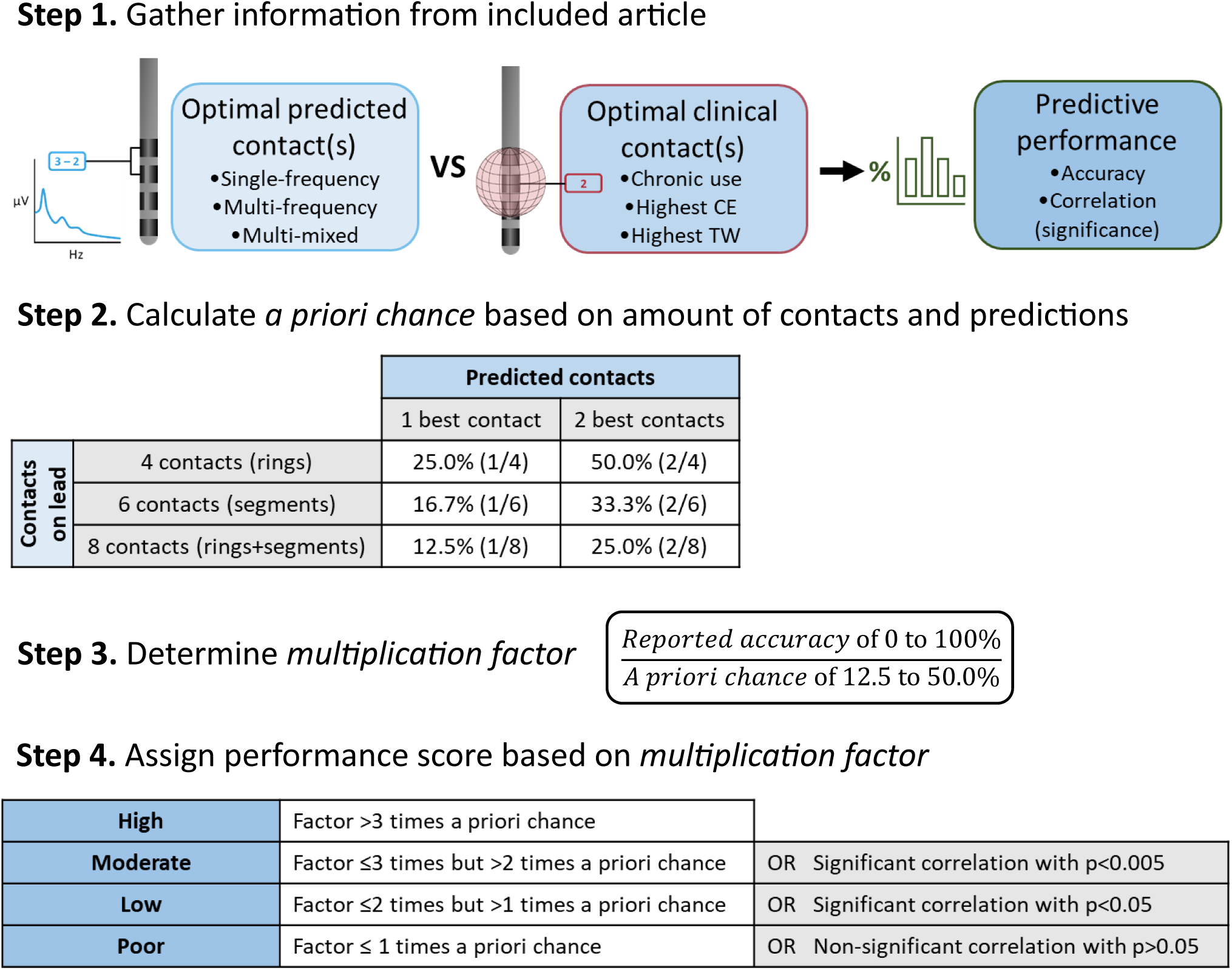
Methodological steps for obtaining custom performance scores based on predictive performance provided by article. In ***Step 1.*** Available information on the predicted optimal contact(s), the optimal clinical contact(s) and the corresponding predictive performance is gathered from the article. In case of a predictive performance in the form of a correlation/significance Step 4 follows. In case of a predictive performance in the form an accuracy Step 2 follows. In ***Step 2***. the *a priori chance* in then calculated based on the amount of contacts on the lead and the amount of predicted contacts. In ***Step 3.*** the *multiplication factor* is then determined by means of dividing the reported accuracy by the a priori *chance*. Finally, in ***Step 4***. the custom performance score is assigned by considering either the *a priori chance* and *multiplication factor*, as determined in Step 2 and 3 OR by considering the significance and corresponding p-value in case a correlation is provided. *For example: an article provides a predictive performance of 50.0% accuracy for a directional lead with 8 contacts and a prediction of the best 2 contacts. The a priori chance for this scenario is 25.0%. The multiplication factor equals 2 (50.0% divided by 25.0%). A factor of 2 times the a priori chance falls in the range of a “Low” custom performance score. Alternatively, if the article had provided the prediction of the single best contact, the a priori chance would decrease to 12.5%, leading to a multiplication factor of 4, and a “High” performance score. In a different scenario where the article provides a correlation of r = 0.5 with a p-value < 0.001, the direct conclusion is a “Moderate” custom performance score.*

Results were categorised in four groups to provide a better overview of the performance scores (table 1): a) single-beta predictions: based on a single beta-oscillation feature; b) single-other-frequency predictions: based on a single frequency feature other than beta; c) multi-frequency predictions: based on features from multiple frequency bands; d) multi-mixed predictions: based on a combination of at least one frequency- and one non-frequency-based feature (i.e. ERNA-, and anatomy: (Euclidean) distance to the image-based sweet-spot). Both forms of multi-feature predictions potentially include the use of machine learning algorithms. Within these four groups, a subdivision was made for measures derived from recordings in monopolar, bipolar, or pseudomonopolar configurations. Here, a pseudomonopolar configuration refers to signals recorded in a bipolar configuration, which is subsequently post-processed into monopolar recordings using custom methodologies.

**Table 1.** Overview of the primary outcome categories with corresponding characteristics and options as well as an overview of the secondary outcome category with its methodological subcategories and corresponding subgroups.

| <u>Primary Outcomes</u> |  |  |
| --- | --- | --- |
| Groups | Characteristics of Group | Options for Characteristics |
| Single-beta predictions | - Frequency band<br>- Feature characteristics | - Full-beta; high-beta; low-beta.<br>- Peak-power; average-power; power-suppression. |
| Single-other-frequency predictions | - Frequency band<br>- Feature characteristics | - Theta; alpha; gamma; HFO.<br>- Peak-power; average-power. |
| Multi-frequency predictions | - Frequency band<br>- Feature characteristics<br>- Feature combination method | - Theta; alpha; beta; gamma; HFO.<br>- Peak-power; average-power.<br>- Ratio; machine learning. |
| Multi-mixed predictions | - Frequency band<br>- Non-frequency features<br>- Feature characteristics<br>- Feature combination method | - Theta; alpha; beta; gamma; HFO.<br>- ERNA; anatomy.<br>- Peak-power; average-power; (Euclidian) distance to image-based sweet-spot.<br>- Combined ranking; machine learning. |
| <u>Secondary Outcomes</u> |  |  |
| Group | Methodological Subcategories | Subcategory Subgroups |
| Single-beta predictions | - Recording system<br>- Recording electrodes<br>- Recording lead<br>- Recording mode<br>- Beta feature<br>- Beta (sub)band<br>- Timing of recording | - Externalised; internalised.<br>- Ring; segments; combination.<br>- Traditional; directional.<br>- Monopolar; pseudomonopolar; bipolar.<br>- Beta-peak; average-beta; beta-suppression.<br>- Full-beta; low-beta; high-beta |
|  | - Time between recording and contact choice<br>- Timing of contact choice<br>- Contact choice method | - Intra-op.; <4 months post-op.; ≥4 months post-op.<br>- ≤48 hour; >48 hours & <4 months; ≥4 months.<br>- ≤48 hours post-op.; >48 hours & <4 months post-op.; ≥4 months post-op.<br>- Chronic stimulation contact; Highest TW/CE contact. |

*Abbreviations: HFO: high frequency oscillations; ERNA: evoked resonant neural activity; intra-op.: intra-operatively; post-op.: post-operatively; TW: contact with largest therapeutic window (difference in stimulation current between effect threshold and side-effect threshold) used as reference for prediction; CE: contact with highest clinical efficiency (stimulation induced improvement in contralateral MDS-UPRS-III (sub)scores OFF-medication) used as reference for prediction; MDS-UPDRS-III: Movement Disorder Society Unified Parkinson's Disease Rating Scale part three; HFO: High frequency oscillation.*

For the secondary outcomes, potential clinically relevant differences in methodology were evaluated for the performance scores obtained by single-beta feature predictions. To do so, performance scores were grouped according to methodological subcategories (table 1). The distribution of performance scores across each subgroup of the secondary outcome subcategories was determined as a percentage per performance score across each subgroup (e.g. subcategory: recording device; subgroup 1: internal recording device; frequency of performance classes: 0% poor, 10% low, 80% moderate, 10% high versus subgroup 2: external recording device, frequency of performance classes: 0% poor, 5% low, 70% moderate, 25% high). Differences between subgroups per subcategory were evaluated through a Pearson Chi-Squared test, where a two-sided p-value < 0.05 was considered significant.

## Results

### Article selection

A total of 418 unique articles were initially identified. After application of the predetermined in- and exclusion criteria 25 articles were included and reviewed (figure 3).

**Figure 3.**
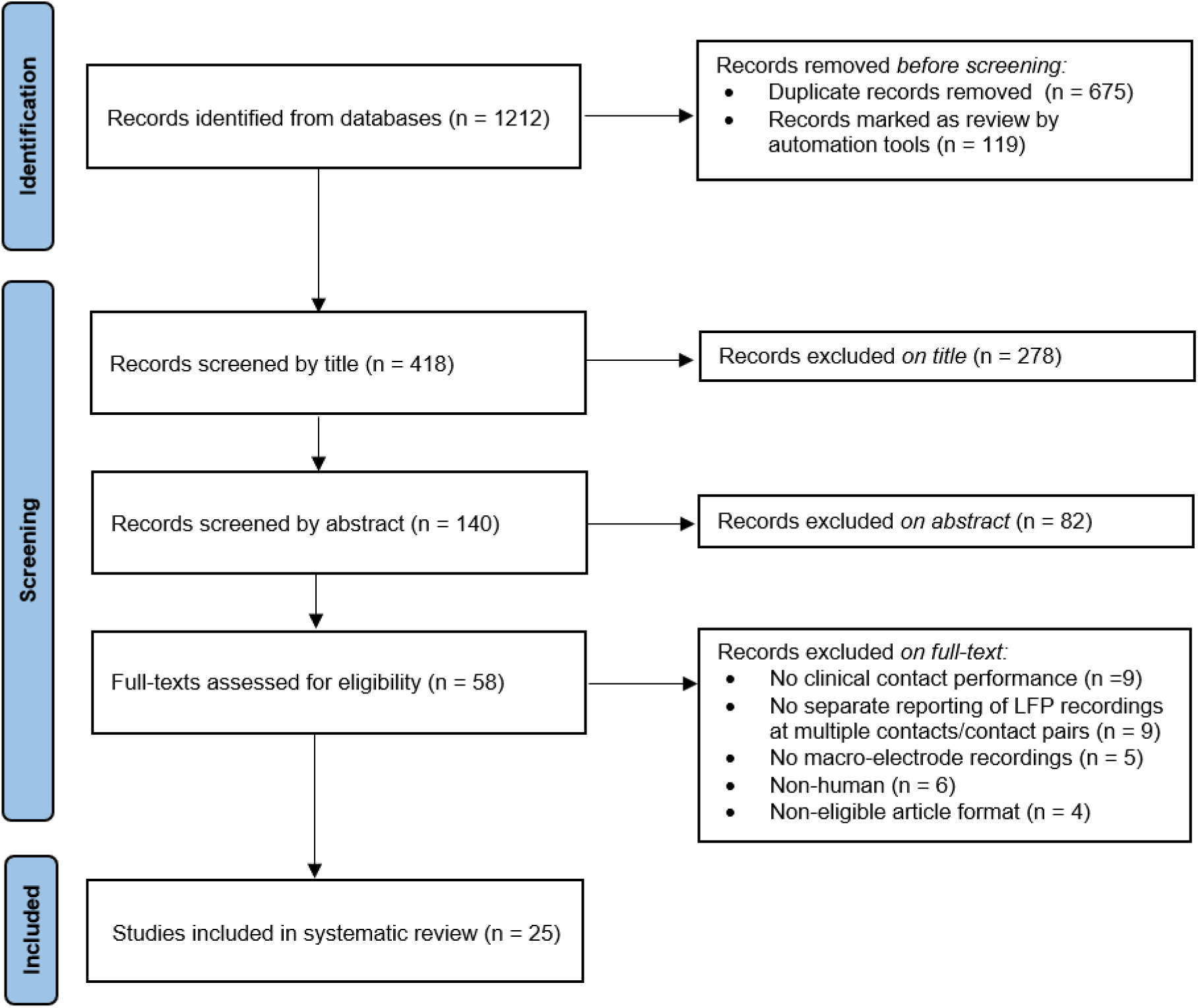
Flow diagram of the study in- and exclusion steps.

### Quality assessment

The median quality score was 13 (range 9-17), supplementary table 1 (Supplementary file 2). A summary of the quality assessment for each study is provided in tables 2, 3 and 4. The included studies have a large variety of recording procedures (e.g. OFF/ON medication, clinician blinding, etc.) and processing methods (e.g. artefact extraction/noise reduction, normalisation, etc.).

**Table 2.** Prediction of the single optimal stimulation contact based on beta-frequency oscillations alone compared to the clinical stimulation contact.

| Article | Prediction method | Outcome | Number of contacts | Score | Number of STN | Medication state | Blinding clinician | JB1 score (Y/N/U) |
| --- | --- | --- | --- | --- | --- | --- | --- | --- |
| <u>Monopolar recordings</u> |  |  |  |  |  |  |  |  |
| [23] | Predictive accuracy β-peak | 59.0% | 8 | high | 28 | U | U | 12/4/2 |
| [32] | Predictive accuracy β-peak | 63% (CE) | 6 | high | 19 | OFF | Y | 12/5/1 |
| [31]<sup>†</sup> | Correlation high-β-peak | $r=-0.452$, $p<0.001$ | 4 | moderate | 50 | U | U | 13/3/2 |
| [37] | Agreement β-peak | 73.3% | 4# | moderate | 45 | OFF | Y | 13/4/1 |
| [33] | Agreement β-peak | 54.2% | 4 | moderate | 24 | U | Y | 13/3/2 |
| [21]<sup>†</sup> | Predictive accuracy low-β-peak | 47.0% (TW) | 6 | moderate | 27 | OFF | Y | 17/1/0 |
| [21]<sup>†</sup> | Predictive accuracy low-β-peak | 30.0% (TW) | 8 | moderate | 27 | OFF | Y | 17/1/0 |
| [20]<sup>†</sup> | Predictive accuracy average-β-power | 50.0% | 4 | low | 92 | U | Y | 12/5/1 |
| [17]<sup>†</sup> | Agreement average-β-power | 50.0% | 4 | low | 28 | U | Y | 12/4/2 |
| [25] | Correlation β-peak | $r^2=0.36$, $p=0.02$ (CE) | 4 | low | 12 | OFF | U | 14/3/1 |
| [15]<sup>†</sup> | Predictive accuracy low-β-peak | 66.7% (TW) | 3 | low | 4 | OFF | U | 11/5/2 |
| [15]<sup>†</sup> | Predictive accuracy β-peak | 58.3% (TW) | 3 | low | 4 | OFF | U | 11/5/2 |
| [15]<sup>†</sup> | Predictive accuracy high-β-peak | 41.7% (TW) | 3 | low | 4 | OFF | U | 11/5/2 |
| [31]<sup>†</sup> | Correlation low-β-peak | $r=-0.067$, $p=0.645$ | 4 | poor | 50 | U | U | 13/3/2 |
| <u>Pseudomonopolar recordings</u> (*bipolar LFP recordings transformed to monopolar configuration using custom technique*) |  |  |  |  |  |  |  |  |
| [34]<sup>†</sup> | Predictive accuracy β-suppression | 50.0% | 6 | moderate | 32 | OFF | Y | 15/2/1 |
| [34]<sup>†</sup> | Predictive accuracy β-peak | 37.5% | 6 | moderate | 32 | OFF | Y | 15/2/1 |
| [30]<sup>†</sup> | Predictive accuracy β-peak | 71.0% (CE) | 3 | moderate | 14 | OFF | Y | 16/1/1 |
| [30]<sup>†</sup> | Predictive accuracy β-peak | 50.0% (CE) | 4 | low | 14 | OFF | Y | 16/1/1 |
| [27] | Agreement β-peak | 33.0% | 4 | low | 9 | OFF | Y | 15/2/1 |
| <u>Bipolar recordings</u> |  |  |  |  |  |  |  |  |
| [29]<sup>†</sup> | Predictive accuracy low-β-suppression | 40.0% | 8 | high | 13 | OFF | Y | 16/1/1 |
| [35] | Correlation low-/high-β-peak | $r=-0.45$, $p=0.0001$ | 6 | moderate | 28 | OFF | Y | 15/2/1 |
| [22]<sup>†</sup> | Difference z-score low-β-power vs. inactive | $p=0.008$ | 4 | low | 95 | OFF | U | 14/3/1 |
| [22]<sup>†</sup> | Difference z-score β-power vs. inactive | $p=0.01$ | 4 | low | 95 | OFF | U | 14/3/1 |
| [11] | Correlation average-β-power change | $r=0.35$, $p=0.01$ | 4 | low | 54 | OFF | Y | 12/5/1 |
| [24] | Agreement β-peak | 47.0% | 4* | low | 19 | OFF | U | 10/7/1 |
| [5] | Difference low-β-peak vs. empirical | p > 0.05 (CE) | 3 | low | 16 | OFF | Y | 11/6/1 |
| [29]<sup>†</sup> | Predictive accuracy high-β-suppression | 25.0% | 8 | low | 13 | OFF | Y | 16/1/1 |
| [26] | Agreement β-peak | 90.0% | 4* | low | 10 | OFF | Y | 15/2/1 |
| [28] | Agreement β-peak | 87.5% | 4* | low | 8 | ON | U | 9/6/1<sup>†</sup> |
| [1] | Agreement β-peak | 100.0% | 4*# | low | 4 | OFF | Y | 14/3/1 |
| [22]<sup>†</sup> | Difference z-score high-β-power vs. inactive | p=0.062 | 4 | poor | 95 | OFF | U | 14/3/1 |

Performance score: High: >3 times *a priori* chance; moderate: $\leq 3$ times but >2 times *a priori* chance OR significant correlation with $p < 0.005$ ; low: $\leq 2$ times but > 1 time *a priori* chance OR significant correlation with $p < 0.05$ ; poor: $\leq 1$ time *a priori* chance OR non-significant correlation with $p > 0.05$ ; \*: both contacts from bipolar recording configuration considered as prediction; #: both contacts from bipolar stimulation configuration used as reference for predictions (supplementary results, Supplementary file 1); <sup>†</sup>: 2 JBI questions were not applicable; <sup>‡</sup>: multiple outcomes presented in the table. Abbreviations: Y: Yes; N: No; U: Unknown/Unclear; STN: subthalamic nucleus; TW: contact with largest therapeutic window (difference in stimulation current between effect threshold and side-effect threshold) used as reference for prediction; CE: contact with highest clinical efficiency (stimulation induced improvement in contralateral MDS-UPDRS-III (sub)scores OFF-medication) used as reference for prediction; MDS-UPDRS-III: Movement Disorder Society Unified Parkinson's Disease Rating Scale part three; HFO: High frequency oscillation.

**Table 3.** Prediction of the single optimal stimulation contact based on non-beta-frequency oscillations compared to the clinical stimulation contact.

| Article | Prediction methods | Outcome | Number of contacts | Score | Number of STN | Medication state | Clinician blinding | JB1 score (Y/N/U) |
| --- | --- | --- | --- | --- | --- | --- | --- | --- |
| <u>Monopolar recordings</u> |  |  |  |  |  |  |  |  |
| [33]<sup>‡</sup> | Agreement HFO-peak | 70.8% | 4 | moderate | 24 | U | Y | 13/3/2 |
| [17] | Agreement average-HFO-power | 36.0% | 4 | low | 28 | U | Y | 12/4/2 |
| [33]<sup>‡</sup> | Agreement $\theta$+α-peak | 25.0% | 4 | poor | 40 | U | Y | 13/3/2 |
| [31]<sup>‡</sup> | Correlation θ-peak | $r=-0.039$, $p=0.786$ (CE) | 4 | poor | 50 | U | Y | 13/3/2 |
| [31]<sup>‡</sup> | Correlation α-peak | $r=-0.131$, $p=0.366$ (CE) | 4 | poor | 50 | U | Y | 13/3/2 |
| [31]<sup>‡</sup> | Correlation low-γ-peak | $r=-0.110$, $p=0.446$ (CE) | 4 | poor | 50 | U | Y | 13/3/2 |
| <u>Bipolar recordings</u> |  |  |  |  |  |  |  |  |
| [1]<sup>‡</sup> | Agreement γ-peak | 100.0% | 4*# | low | 4 | OFF | Y | 14/3/2 |
| [22] | Difference z-score α-power vs. inactive | $p=0.112$ | 4 | poor | 95 | OFF | U | 14/3/1 |
| [35] | Correlation γ-peak | $r=0.17$, $p=0.43$ | 6 | poor | 28 | OFF | Y | 15/2/1 |
| [36] | Z-score γ-power > empirical | $p>0.05$ (CE) | 4 | poor | 20 | OFF | Y | 9/6/3 |
| [1]<sup>‡</sup> | Agreement θ-peak | 0.0% | 4*# | poor | 4 | OFF | Y | 14/3/2 |
| [1]<sup>‡</sup> | Agreement α-peak | 0.0% | 4*# | poor | 4 | OFF | Y | 14/3/2 |

Performance score: High: >3 times *a priori* chance; moderate: $\leq 3$ times but >2 times *a priori* chance OR significant correlation with $p<0.005$ ; low: $\leq 2$ times but > 1 time *a priori* chance OR significant correlation with $p<0.05$ ; poor: $\leq 1$ time *a priori* chance OR non-significant correlation with $p>0.05$ ; \*: both contacts from bipolar recording configuration considered as prediction; #: both contacts from bipolar stimulation configuration used as reference for predictions (supplementary results, Supplementary file 1); <sup>‡</sup>: multiple outcomes presented in the table. Abbreviations: Y: Yes; N: No; U: Unknown/Unclear; STN: subthalamic nucleus; HFO: High frequency oscillation; CE: contact with highest clinical efficiency (stimulation induced improvement in contralateral MDS-UPRS-III (sub)scores OFF-medication) used as reference for prediction; MDS-UPDRS-III: Movement Disorder Society Unified Parkinson's Disease Rating Scale part three.

**Table 4.** Prediction of the single optimal stimulation contact based on beta-frequency oscillations in combination with other (non-)frequency features compared to the clinical stimulation contact.

| Article | Prediction methods | Outcome | Number of contacts | Score | Number of STN | Medication state | Clinician blinding | JB1 score (Y/N/U) |
| --- | --- | --- | --- | --- | --- | --- | --- | --- |
| Multiple frequency features only |  |  |  |  |  |  |  |  |
| <u>Monopolar recordings</u> |  |  |  |  |  |  |  |  |
| [21]<sup>‡</sup> | LASSO ($\alpha$, low/high-β, (fast)-γ, HFO) | 37.0% (TW) | 6 | moderate | 27 | OFF | Y | 17/1/0 |
| [21]<sup>‡</sup> | LASSO ($\alpha$, low/high-β, (fast)-γ, HFO) | 30.0% (TW) | 8 | moderate | 27 | OFF | Y | 17/1/0 |
| <u>Bipolar recordings</u> |  |  |  |  |  |  |  |  |
| [16] | SVM (θ- and low-β-power) | 91.0% | 4 | high | 28 | OFF | U | 11/5/2 |
| [36] | $\beta/\alpha$-ratio z-score vs. empirical | $p < 0.001$ (CE) | 4 | moderate | 20 | OFF | U | 9/6/3 |
| Multiple frequency and non-frequency features |  |  |  |  |  |  |  |  |
| <u>Monopolar recordings</u> |  |  |  |  |  |  |  |  |
| [20]<sup>‡</sup> | RF (average-ERNA/-β+anatomy) | 80.0% | 4 | high | 92 | U | Y | 12/5/1 |
| [17]<sup>‡</sup> | Ranking (average-ERNA, -β, -HFO) | $r^2=0.61$, $p<0.001$ (CE) | 4 | moderate | 28 | U | Y | 12/4/2 |

Performance score: High: $>3$ times *a priori* chance; moderate: $\leq 3$ times but $>2$ times *a priori* chance OR significant correlation with $p<0.005$ ; low: $\leq 2$ times but $>1$ time *a priori* chance OR significant correlation with $p<0.05$ ; poor: $\leq 1$ time *a priori* chance OR non-significant correlation with $p>0.05$ ; <sup>‡</sup>: multiple outcomes presented in the table. Abbreviations: Y: Yes; N: No; U: Unknown/Unclear; STN: subthalamic nucleus; LASSO: Least Absolute Shrinkage and Selection Operator regression; HFO: High frequency oscillation; SVM: support vector machine; RF: Random Forest; ERNA: evoked resonant neural activity; anatomy: Euclidean distance to the image-based sweet-spot; TW: contact with highest therapeutic window (difference in stimulation current between effect threshold and side-effect threshold) used as reference for prediction; CE: contact with highest clinical efficiency (stimulation induced improvement in contralateral MDS-UPRS-III (sub)scores OFF-medication) used as reference for prediction; MDS-UPDRS-III: Movement Disorder Society Unified Parkinson's Disease Rating Scale part three.

Among the 25 included studies, 5 studies concerned retrospective research [16, 20–23], 11 studies analysed data from ten or less patients [1, 5, 13, 15, 24–30], 17 articles originated from the work of six independent research groups (group #1: [11, 13, 15, 21, 31, 32]; group #2: [17, 20, 33]; group #3: [1, 16]; group #4: [5, 24]; group #5: [22, 34]; group #6: [29, 30]). Some articles included data collected from partially overlapping patient populations [11, 13, 29, 30].

### Study characteristics

This systematic review encompassed a total of 763 STN placed leads, with a median of 27 STN placed leads per study (IQR: 27). A comprehensive summary of the study characteristics, methods and outcomes of the included articles is provided in supplementary table 2 (Supplementary file 2).

Recordings were collected by means of an externalised device (17 studies; 624 STN) or via an implantable pulse generator (IPG) (8 studies; 139 STN); using traditional leads (16 studies: 582 STN), directional leads (8 studies: 153 STN) or both (1 study: 1 STN traditional and 27 STN directional). The choice of the *clinical stimulation contact* was based on either MPR (13 studies; 278 STN), the chronic stimulation settings (11 studies: 466 STN) or both (1 study: 17 STN MPR and 2 STN chronic stimulation settings). The median time of the *clinical stimulation contact* choice ranged from several minutes to 19 months after primary lead placement. In general, the LFP recordings were performed several minutes up to 6 months after initial lead placement. In three studies (19 STN) recordings were performed during IPG replacements.

### Primary outcomes

When considering single contact predictions based on LFP recordings in the beta-frequency band, the large majority of predictions (94%) achieved either a predictive accuracy larger than the *a priori* chance or a significant correlation with the *clinical stimulation contact* (table 2, figure 4). Only two outcomes did not achieve significance (poor performance score): the first, (r=-0.067, p=0.645) used monopolar recordings and a low-beta-peak feature [31]; the second, (p=0.062) used bipolar recordings and a z-score for high-beta-power [22]; notably, in both studies other considered beta-related features were significantly correlated. For 39% a predictive accuracy of two times the *a priori* chance or more (moderate or high performance score) was achieved. In this category a high amount (6 or 8) of contact points was more often used than a low (3 or 4) amount of contact points (67% vs. 33%, respectively).

**Figure 4.**
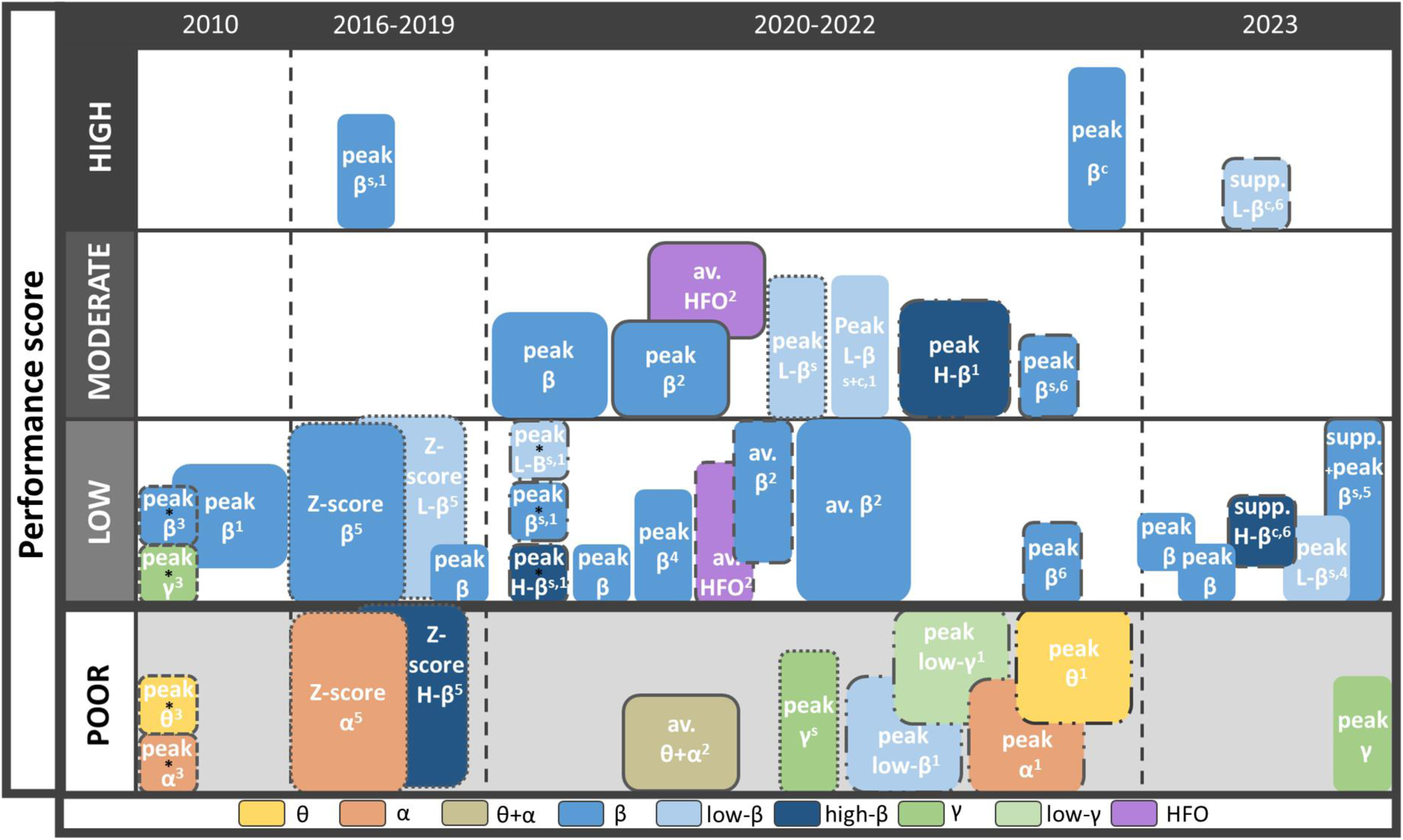
Overview of outcomes according to the used LFP feature for studies with a single feature approach. The figure shows the predictive value of a specific LFP frequency feature (colour-coded) in predicting the single most optimal stimulation contact for chronic stimulation. The figure demonstrates that positive outcomes were solely achieved using beta-related features with the exception of two studies using HFO and one using gamma. Studies using other LFP features reported almost exclusively no correlation. Performance score: High: >3 times *a priori* chance; moderate: ≤3 times but >2 times *a priori* chance OR significant correlation with p<0.005; low: ≤2 times but > 1 time *a priori* chance OR significant correlation with p<0.05; poor: ≤ 1 time *a priori* chance OR non-significant correlation with p>0.05. Corresponding borders of the boxes indicate results coming from the same study; The size of the box is proportional to the amount of STN nuclei reported; *studies including ≤ 4 subthalamic 18 nuclei; β^s^: segment recordings; β^c^: combination of ring-level and segment recordings; β^#^: research group number. *<u>Abbreviations:</u> av.: average; supp.: suppression; L-/H-β: low/high-β; HFO: high frequency osscillations*.

Using the best two (and not single) *clinical stimulation contacts* as clinical outcome never resulted in poor performance scores (Supplementary table 3, Supplementary file 2).

The results for predictions based on single frequency features other than beta-band features achieved low to moderate scores in 25% of the outcomes (table 3, figure 4). Only studies evaluating HFO-based features consistently resulted in low or moderate scores [17, 33]. Gamma-band feature predictions achieved a low performance in a single study across 4 STN, however, all other gamma-based predictions, including two larger studies, achieved a poor performance [1, 31, 35, 36]. Predictions based on theta, alpha, or both feature-bands combined always resulted in poor performance scores. Interestingly, combining multiple features, including frequency and/or non-frequency features, for instance, by means of machine learning, achieved a predictive accuracy of more than two times the *a priori chance* or a significant correlation with p<0.005 in all cases (table 4, figure 5). Therefore, combinations of multiple (frequency-)features obtained more often moderate or high performance scores compared to single beta-based features (100% vs. 39%, respectively).

**Figure 5.**
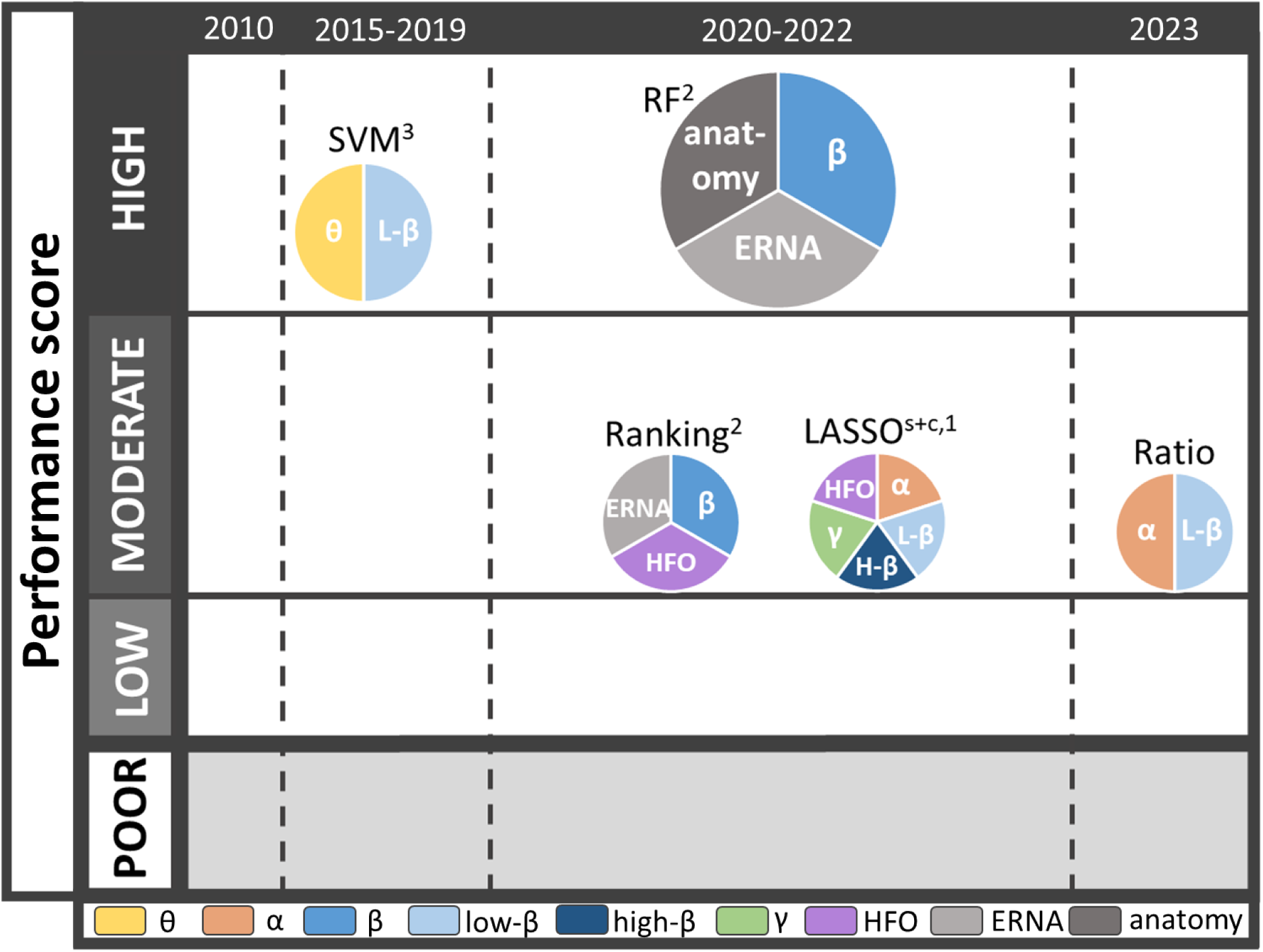
Overview of outcomes according to the used LFP feature for studies with a multi-feature approach. The figure shows the predictive value of a specific prediction technique and combination of (non-)frequency features (colour-coded) in predicting the best contact point for chronic stimulation. Studies using multi-feature predictions always reported (strongly) positive correlations. Performance score: High: >3 times *a priori* chance; moderate: ≤3 times but >2 times *a priori* chance OR significant correlation with p<0.005; low: ≤2 times but > 1 time *a priori* chance OR significant correlation with p<0.05; poor: ≤ 1 time *a priori* chance OR non-significant correlation with p>0.05. The size of the box is proportional to the amount of STN nuclei reported; β^s^: segment recordings; β^c^: combination of ring-level and segment recordings; β^#^: research group number. *<u>Abbreviations</u>: L-/H-β: low-/high-β; ERNA: evoked resonant neural activity; anatomy: Euclidean distance to the image-based sweet-spot; HFO: high frequency oscillations; SVM: support vector machine; RF: random forest; LASSO: least absolute shrinkage and selection operator*.

### Secondary outcomes

Potential clinically relevant differences in methodology were further evaluated using subcategory comparisons for all single beta-based predictions (table 5). Significant differences between subgroups according to the Pearson Chi-squared test were found in only one subgroup, the type of electrode used for recording (χ^2^: 14.825, p = 0.022). The combination of ring-levels and segments (all 8 contacts on a directional lead) results in a higher occurrence of high performance scores, followed by (6) segment-only recordings with a higher frequency of moderate performance scores and finally (4) ring-only recordings with a higher occurrence of low performance scores. However, when comparing the type of lead by itself (directional or traditional) by considering the same number of used electrodes (3 segments only or 4 levels only), no significant difference between directional leads and traditional leads was found.

**Table 5.**
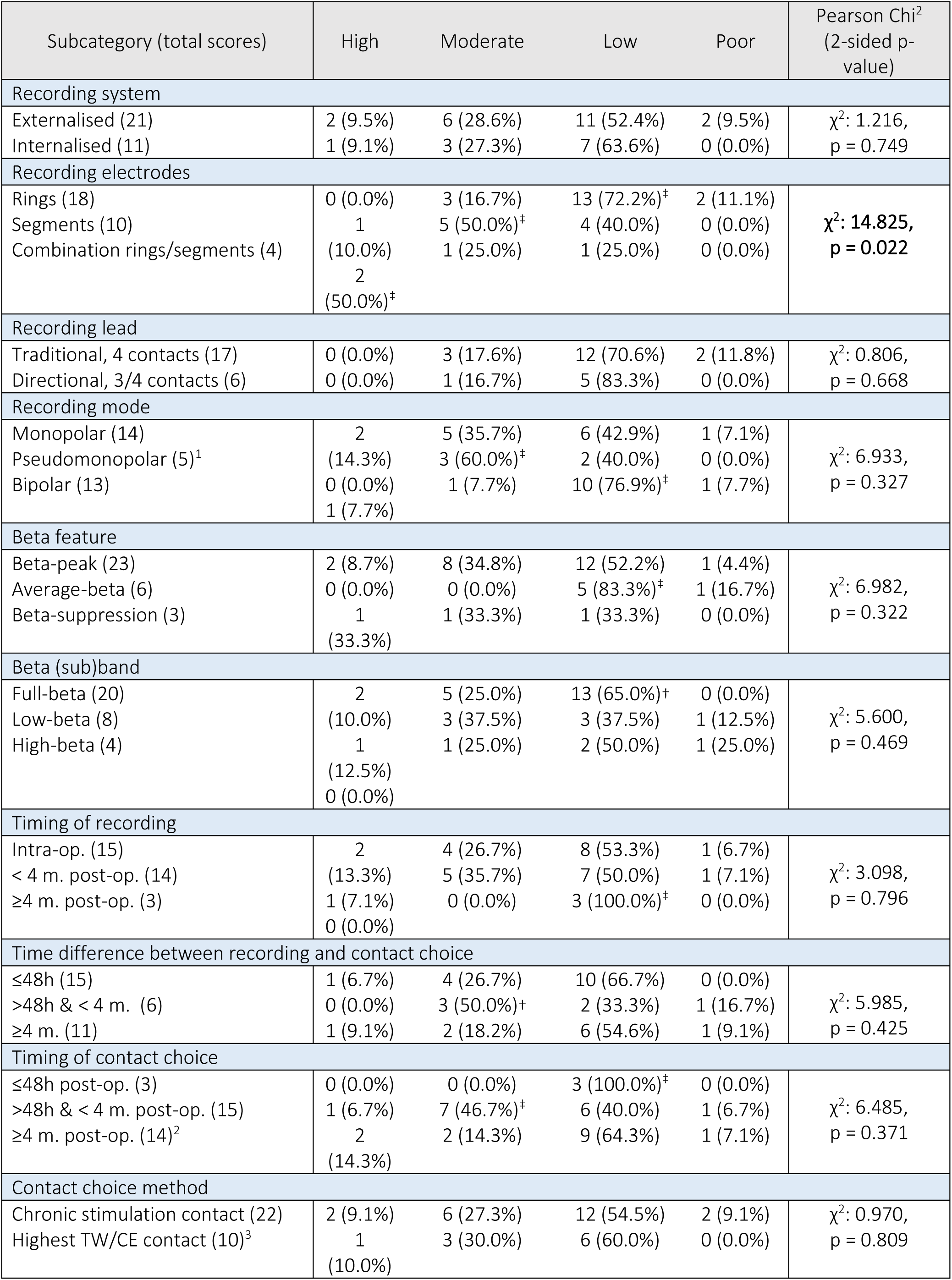
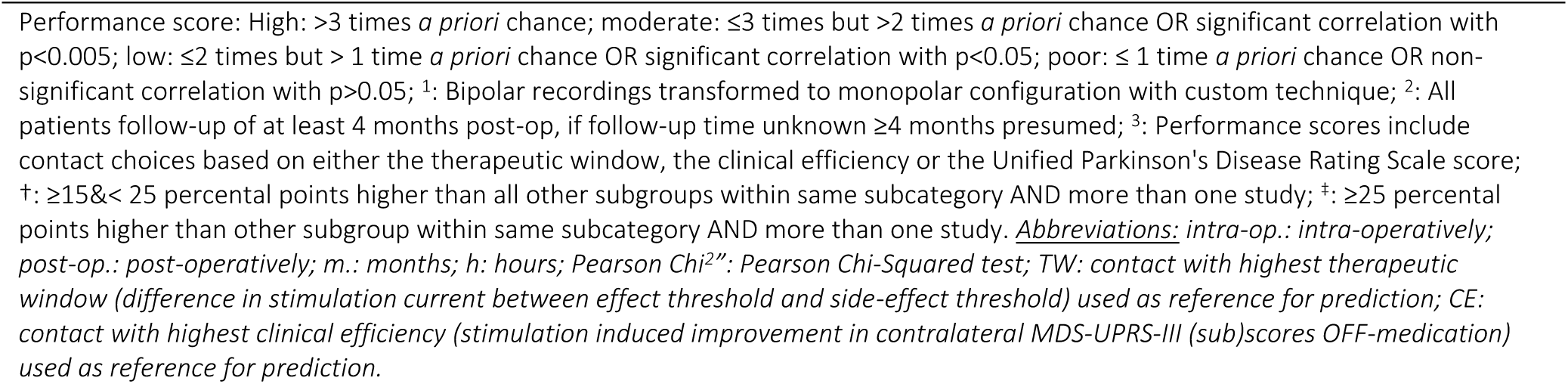
Scores for methodological subcategories in studies using one contact prediction based on beta-oscillations alone.

Several studies also performed a variety of clinical comparisons between LFP-based and empirical *clinical stimulation contact* choices; see supplementary results for an overview (Supplementary file 1). Six studies showed that LFP-based programming can achieve a clinical efficiency that is similar to empirical programming, whilst requiring less programming iterations, programming time, and stimulation voltage [5, 11, 27, 29, 30, 36]. Two studies looked at the effect of general anaesthesia on the predictive value of several neurophysiological factors, however, results remained inconclusive [23, 33]. Five studies evaluated the effect of active or passive movement on the predictive value of several neurophysiological factors: although various differences across the predictive value of different neurophysiological factors were presented, no ideal moving- or resting-state feature(s) could be identified [1, 15, 21, 26, 34].

## Discussion

In this systematic review we provide an overview of different methods used in the current literature to identify LFP-based physiomarkers able to predict *clinical stimulation contact* choices for DBS in PD.

The results indicate that 1) Beta-frequency information is most often used and achieves positive results (low, moderate or high performance scores) in 94% of the outcomes, although methods and results vary largely between studies; 2) Frequency features from other frequency bands provide positive scores in only 25% of the outcomes, all of which concerning either HFO or, less often, gamma frequency features; and 3) Combining various (non-) frequency features shows moderate or high performance scores more often than single beta-band features alone (100% versus 39%, respectively).

### Beta-frequency

In general, this review confirms that predictions based on beta-frequency, including low, high, or total beta, provide positive prediction scores in the majority of the outcomes (29 out of 31). Whether prediction scores of less than 3 times the *a priori* chance (i.e. less than a high performance score) are acceptable in clinical practice will depend on the intended use of the feature. Currently, clinical choice is still to be considered the gold standard, but the use of LFP can be a useful support to complement clinical observations in selected cases or in order to reduce the search window.

In some recent articles the predictive capacity of the low- and high-beta sub-bands are compared. Previous research has shown that these sub-bands may have different physiological meaning and provide different predictive capacity and patterns of association to PD motor symptoms [31, 38–40].

Consequently, we evaluated whether low- and high-beta had a difference in performance when predicting the best stimulation spot, however, current results are not conclusive in that sense.

Earlier articles mainly focussed on the location of the (beta-)frequency peaks, whereas more recent work also addressed the location of the highest average beta-power or the location with the largest stimulation-induced beta-suppression. The rationale for using these additional beta-related features is multifold: an average is less susceptible to noise and/or artefacts than single peaks, and might therefore produce more robust results, and the use of stimulation-induced beta-suppression may serve as a more direct confirmation of clinical efficacy [5, 29]. From our data analysis however, we could not confirm the superiority of these beta-features. It should be noticed that to date, only few articles have evaluated these non-peak beta-features, probably making this comparison less reliable.

### Other frequencies

In the existing literature features related to the theta-, alpha-, gamma- or HFO-band provided poorer results than beta-band features (poor performance in 9/12 outcomes), when considered as single predictors. Several reasons could explain this finding. For example, these frequencies may be related to specific symptoms which are usually not major determinants in the initial choice of the *clinical stimulation contact*, such as dyskinesia, tremor, or othe motor- or non-symptoms. Nonetheless, these frequencies might still be relevant in a chronic setting or in specific subsets of patients, and therefore should not be dismissed based on the limited data available.

### Combination of features

All five studies reporting on the use of combined features achieved moderate to high scores. Two of these studies also individually compared results to predictions based on beta-frequency features alone and showed that combinations of various (non-)frequency features provided superior predictions [16, 20]. The finding that a combination of frequency features can outperform beta-based predictions alone is interesting and should be further explored. Due to the heterogeneity of clinical phenotypes and disease stages of PD patients undergoing DBS, it could be hypothesised that not all aspects of PD can be captured by beta-frequency alone and that taking different features into account might produce better results in individual patients [41]. Considering the wide variation in included features and (machine learning) models used so far, and the potentially even wider variety of methods not yet explored in the available literature, it remains unclear which method and features could provide the best results.

### Methodological aspects

The validity of LFP-based predictions of the *clinical stimulation contact* is possibly influenced by numerous methodological factors such as study size, recording modality, recording instruments, pre-processing method, timing of recording, used reference method and the applied outcome measure. We attempted at identifying factors that might influence the predictive value of LFPs, and found that only the type of recording contact (ring-level vs. segment vs. combination of ring-levels and segments) used for recording seemed to influence the results. Stimulation level predictions were found to be more accurate for ring-levels in segmented electrodes than for cylindrical ring electrodes alone. This could potentially be very relevant, as directional leads require longer clinical evaluations during MPR, and thus a semi-automated *clinical stimulation contact* selection would be most welcome [5, 15, 21, 23, 32, 34]. However, we acknowledge that this difference may be inherent to our analysis method which takes the *a priori* chance into account and thus assigns a higer value to predictions based on a larger number of contacts. Another possible explanation may lie in the fact that some of the directional leads used are those of the last generation, expressely designed with features that facilitate optimal recordings with respect to the tradional leads. Nonetheless, in this research we did not find a significant difference in accuracy between predictions performed with directional or traditional leads.

### Other methodological aspects relevant for future research

No other methodological aspect was significantly related to the prediction efficacy. It is important to notice that sub-analysis was only possible for studies reporting results on beta-related features because these were present in larger numbers. Most of these studies yield positive results and used a large variability of methods, which could make it difficult to identify potential small effects of methodological factors. On the other end we cannot exclude that some of these methodological factors might be more relevant when investigating other LFP features.

One important aspect could be the use of monopolar recordings with respect to bipolar recordings, because the latter require a translation to a single stimulation contact which still remains a complex matter [30].

Growing evidence suggests that the features of LFP recordings can change over time, especially in the early post-operative period, probably due to the stun-effect [42] or other surgery-related factors (such as patient fatigue, possible sedation residues). While most of the earlier studies were bound to recordings during surgery or in the immediate postoperative period, due to de inavailability of implantable systems recording LFPs, a large number of the reviewed studies (27 out of 32 outcomes) analysed LFP recordings and clinical contact choices performed later on, which is probably a more robust design to consider for future studies.

Although recordings from externalised systems bear the possibility of more powerful analysis techniques (e.g. higher sampling frequency), in this review they did not result in higher accuracies.

### Strengths and Limitations

In this review we chose to focus on the clinical contact choice based on LFPs, an aspect which is highly relevant for the current clinical practice. Since the introduction on the market of the new implantable DBS systems capable of LFP recordings, an increasing number of clinics are presently using or considering the integration of LFP recordings into their clinical stimulation adjustment protocols with the aim of reducing time and resources needed for this complex process. Consequently, the findings of this review lay the groundwork for future research and, by extension, future clinical protocols in DBS programming.

Here, we systematically reviewed the use of various frequency (and non-frequency) features, models and feature combinations from a vast amount of studies from multiple databases. Furthermore, various methodological issues which are crucial for the design of future studies, such as the modality, pre-processing and timing of LFP recordings, were reviewed. Considering the variability of hardware and methods used in the literature, we adopted a novel more objective classification of outcomes in scoring classes, which takes into account the *a priori* chance of prediction. This method helped eliminate variations in performance due to methodological differences. Nonetheless, the performance score also comes with its limitations; as it allows for a high variation in outcome methods, the performance score may have introduced bias in favour of specific outcomes. Additionally, due to the variability in the available studies, each scoring class includes a range of prediction methods, reference methods, and study population sizes.

Importantly, not all of the reviewed articles implemented normalisation, noise reduction, and/or artefact extraction. Although these factors were considered in the quality assessment, it remained challenging to evaluate if and how they influenced the results. There was also a slight variation in the frequency band definitions across studies, and a high heterogeneity was present in the methods for recording LFPs. Furthermore, a large part of the included studies were conducted across a small number of centres, and for six research groups more than one article was included in this review, which may have led to bias. Nonetheless, as this is a new technique and these few centres have built up expertise regarding the subject, this may have also increased the reliability of the results. For the secondary outcomes, an important drawback is that several articles were included multiple times.

Finally, most of the included articles considered the chronic stimulation contact as a reference measure. However, it is important to notice that the chronic stimulation contact choice does not necessarily reflect the contact with the greatest motor improvement, and can be influenced by side-effects, which may not be reflected in LFP recordings. Furthermore, the chronic stimulation contact choice may also be biased by patient or clinician preferences Identifying the most optimal reference contact based on TW or CE as rated during the monopolar review could be considered more objective, however these contacts may not always represent the best option for chronic stimulation (e.g. due to delayed-onset (side)-effects). In a recent study, we showed that in 86% of the cases, there is little difference between contacts chosen during the MPR (TW/CE) and those used at the 6-month to 1-year follow-up (chronic)[43].

### Conclusions and future perspectives

Research focusing on the use of LFPs for determining the optimal *clinical stimulation contact* has increased exponentially since 2010. There is a wide variability of methods used concerning the pre-processing steps; methods for converting bipolar recordings to monopolar predictions; timing for LFP recordings and clinical contact choice; and applied prediction techniques/(machine learning) models: all these factors should be accurately considered within future research.

While earlier articles focussed on the location of the (beta-)frequency peaks alone, more recent work also investigates other frequency features and even dynamic features such as stimulation-induced beta-suppression. Single beta-frequency feature predictions show more promising results than predictions by means of single features from other frequency bands. Future research should focus on systematically comparing the predictive capacity of different beta-(sub-)band features.

Furthermore, in the most recent literature there seems to be a trend toward multi-feature predictions and the use of machine learning methods. Evidence suggests that beta-based predictions may be improved by integrating features from other frequencies or non-frequency features. Future research should focus on the identification of multi-feature combinations and models with the highest predictive capacity for individualised predictions.

Results from this review could serve as the basis for future research and clinical protocols in DBS programming, adding objective LFP-based support to the clinical contact selection process.

## Data Availability

All data produced in the present work are contained in the manuscript or available online in the original publication.

## Acknowledgements

We wish to acknowledge the contribution of J.W. (Jan) Schoones, librarian LUMC, for assisting with the search.

## CRediT Statement

- M. Muller: Conceptualization; Data curation; Formal analysis; Investigation; Methodology; Visualization; Writing – original draft
- M.F.C. van Leeuwen: Formal analysis; Investigation; Writing – original draft
- C.F. Hoffmann: Writing – review & editing
- N.A. van der Gaag: Writing – review & editing
- R. Zutt: Writing – review & editing
- S. van der Gaag: Writing – review & editing
- A.C. Schouten: Conceptualization; Supervision; Writing – review & editing
- M.F. Contarino: Conceptualization; Methodology; Visualization; Supervision; Writing – review & editing

## References

[1] Ince NF, Gupte A, Wichmann T, Ashe J, Henry T, Bebler M, et al. Selection of optimal programming contacts based on local field potential recordings from subthalamic nucleus in patients with Parkinson’s disease. Neurosurgery. 2010;67(2):390–7.

[2] Pintér D, Járdaházi E, Balás I, Harmat M, Makó T, Juhász A, et al. Antiparkinsonian Drug Reduction After Directional Versus Omnidirectional Bilateral Subthalamic Deep Brain Stimulation. Neuromodulation. 2023;26(2):374–81.

[3] Volkmann J, Moro E, Pahwa R. Basic algorithms for the programming of deep brain stimulation in Parkinson’s disease. Mov Disord. 2006;21 Suppl 14:S284–9.

[4] Thompson JA, Lanctin D, Ince NF, Abosch A. Clinical implications of local field potentials for understanding and treating movement disorders. Stereotact Funct Neurosurg. 2014;92(4):251–63.

[5] Binder T, Lange F, Pozzi N, Musacchio T, Daniels C, Odorfer T, et al. Feasibility of local field potential-guided programming for deep brain stimulation in Parkinson’s disease: A comparison with clinical and neuro-imaging guided approaches in a randomized, controlled pilot trial. Brain Stimulation. 2023;16(5):1243–51.

[6] Pollo C, Kaelin-Lang A, Oertel MF, Stieglitz L, Taub E, Fuhr P, et al. Directional deep brain stimulation: an intraoperative double-blind pilot study. Brain. 2014;137(Pt 7):2015–26.

[7] Steigerwald F, Müller L, Johannes S, Matthies C, Volkmann J. Directional deep brain stimulation of the subthalamic nucleus: A pilot study using a novel neurostimulation device. Mov Disord. 2016;31(8):1240–3.

[8] Contarino MF, Bour LJ, Verhagen R, Lourens MA, de Bie RM, van den Munckhof P, et al. Directional steering: A novel approach to deep brain stimulation. Neurology. 2014;83(13):1163–9.

[9] Mestre TA, Lang AE, Okun MS. Factors influencing the outcome of deep brain stimulation: Placebo, nocebo, lessebo, and lesion effects. Movement Disorders. 2016;31(3):290–8.

[10] Kühn AA, Volkmann J. Innovations in deep brain stimulation methodology. Movement Disorders. 2017;32(1):11–9.

[11] Yoshida F, Martinez-Torres I, Pogosyan A, Holl E, Petersen E, Chen CC, et al. Value of subthalamic nucleus local field potentials recordings in predicting stimulation parameters for deep brain stimulation in Parkinson’s disease. J Neurol Neurosurg Psychiatry. 2010;81(8):885–9.

[12] Zaidel A, Spivak A, Grieb B, Bergman H, Israel Z. Subthalamic span of beta oscillations predicts deep brain stimulation efficacy for patients with Parkinson’s disease. Brain. 2010;133(Pt 7):2007–21.

[13] Chen CC, Pogosyan A, Zrinzo LU, Tisch S, Limousin P, Ashkan K, et al. Intra-operative recordings of local field potentials can help localize the subthalamic nucleus in Parkinson’s disease surgery. Exp Neurol. 2006;198(1):214–21.

[14] Holdefer RN, Cohen BA, Greene KA. Intraoperative local field recording for deep brain stimulation in Parkinson’s disease and essential tremor. Mov Disord. 2010;25(13):2067–75.

[15] Nguyen TAK, Schüpbach M, Mercanzini A, Dransart A, Pollo C. Directional Local Field Potentials in the Subthalamic Nucleus During Deep Brain Implantation of Parkinson’s Disease Patients. Front Hum Neurosci. 2020;14:521282.

[16] Connolly AT, Kaemmerer WF, Dani S, Stanslaski SR, Panken E, Johnson MD, et al., editors. Guiding deep brain stimulation contact selection using local field potentials sensed by a chronically implanted device in Parkinson’s disease patients. 2015 7th International IEEE/EMBS Conference on Neural Engineering (NER); 2015: IEEE.

[17] Xu SS, Sinclair NC, Bulluss KJ, Perera T, Lee WL, McDermott HJ, et al. Towards guided and automated programming of subthalamic area stimulation in Parkinson’s disease. Brain Commun. 2022;4(1):fcac003.

[18] Moher D, Liberati A, Tetzlaff J, Altman DG. Preferred reporting items for systematic reviews and meta-analyses: the PRISMA statement. PLoS Med. 2009;6(7):e1000097.

[19] Munn Z, Barker TH, Moola S, Tufanaru C, Stern C, McArthur A, et al. Methodological quality of case series studies: an introduction to the JBI critical appraisal tool. JBI Evid Synth. 2020;18(10):2127–33.

[20] Xu SS, Lee WL, Perera T, Sinclair NC, Bulluss KJ, McDermott HJ, et al. Can brain signals and anatomy refine contact choice for deep brain stimulation in Parkinson’s disease? J Neurol Neurosurg Psychiatry. 2022.

[21] Shah A, Nguyen TK, Peterman K, Khawaldeh S, Debove I, Shah SA, et al. Combining Multimodal Biomarkers to Guide Deep Brain Stimulation Programming in Parkinson Disease. Neuromodulation. 2023;26(2):320–32.

[22] Horn A, Neumann WJ, Degen K, Schneider GH, Kühn AA. Toward an electrophysiological “sweet spot” for deep brain stimulation in the subthalamic nucleus. Hum Brain Mapp. 2017;38(7):3377–90.

[23] di Biase L, Piano C, Bove F, Ricci L, Caminiti ML, Stefani A, et al. Intraoperative Local Field Potential Beta Power and Three-Dimensional Neuroimaging Mapping Predict Long-Term Clinical Response to Deep Brain Stimulation in Parkinson Disease: A Retrospective Study. Neuromodulation. 2023.

[24] Thenaisie Y, Palmisano C, Canessa A, Keulen BJ, Capetian P, Castro Jimenez M, et al. Towards adaptive deep brain stimulation: clinical and technical notes on a novel commercial device for chronic brain sensing. Journal of neural engineering. 2021;13.

[25] Tamir I, Wang D, Chen W, Ostrem JL, Starr PA, de Hemptinne C. Eight cylindrical contact lead recordings in the subthalamic region localize beta oscillations source to the dorsal STN. Neurobiol Dis. 2020;146:105090.

[26] Kochanski RB, Shils J, Verhagen Metman L, Pal G, Sani S. Analysis of Movement-Related Beta Oscillations in the Off-Medication State During Subthalamic Nucleus Deep Brain Stimulation Surgery. J Clin Neurophysiol. 2019;36(1):67–73.

[27] Lewis S, Radcliffe E, Ojemann S, Kramer DR, Hirt L, Case M, et al. Pilot Study to Investigate the Use of In-Clinic Sensing to Identify Optimal Stimulation Parameters for Deep Brain Stimulation Therapy in Parkinson’s Disease. Neuromodulation. 2023.

[28] Swinnen BEKS, Stam MJ, Buijink AWG, de Neeling MGJ, Schuurman PR, de Bie RMA, et al. Employing LFP recording to optimize stimulation location and amplitude in chronic DBS for P k ’ : of-concept pilot study. Deep Brain Stimulation. 2023;2:1–5.

[29] Strelow JN, Dembek TA, Baldermann JC, Andrade P, Fink GR, Visser-Vandewalle V, et al. Low beta-band suppression as a tool for DBS contact selection for akinetic-rigid symptoms in Parkinson’s disease. Parkinsonism Relat Disord. 2023;112:105478.

[30] Strelow JN, Dembek TA, Baldermann JC, Andrade P, Jergas H, Visser-Vandewalle V, et al. Local Field Potential-Guided Contact Selection Using Chronically Implanted Sensing Devices for Deep Brain Stimulation in Parkinson’s Disease. Brain Sci. 2022;12(12).

[31] Chen PL, Chen YC, Tu PH, Liu TC, Chen MC, Wu HT, et al. Subthalamic high-beta oscillation informs the outcome of deep brain stimulation in patients with Parkinson’s disease. Front Hum Neurosci. 2022;16:958521.

[32] Tinkhauser G, Pogosyan A, Debove I, Nowacki A, Shah SA, Seidel K, et al. Directional local field potentials: A tool to optimize deep brain stimulation. Mov Disord. 2018;33(1):159–64.

[33] Sinclair NC, McDermott HJ, Lee WL, Xu SS, Acevedo N, Begg A, et al. Electrically evoked and spontaneous neural activity in the subthalamic nucleus under general anesthesia. J Neurosurg. 2021:1–10.

[34] Busch JL, Kaplan J, Bahners BH, Roediger J, Faust K, Schneider GH, et al. Local Field Potentials Predict Motor Performance in Deep Brain Stimulation for Parkinson’s Disease. Movement Disorders. 2023.

[35] Fernandez-Garcia C, Monje MHG, Gomez-Mayordomo V, Foffani G, Herranz R, Catalan MJ, et al. Long-term directional deep brain stimulation: Monopolar review vs. local field potential guided programming. Brain Stimulation. 2022;15(3):727–36.

[36] Dong W, Qiu C, Chang L, Sun J, Yan J, Luo B, et al. The guiding effect of local field potential during deep brain stimulation surgery for programming in Parkinson’s disease patients. CNS Neuroscience and Therapeutics. 2023.

[37] Milosevic L, Scherer M, Cebi I, Guggenberger R, Machetanz K, Naros G, et al. Online Mapping With the Deep Brain Stimulation Lead: A Novel Targeting Tool in Parkinson’s Disease. Mov Disord. 2020;35(9):1574–86.

[38] Morelli N, Summers RLS. Association of subthalamic beta frequency sub-bands to symptom severity in patients with Parkinson’s disease: A systematic review. Parkinsonism & Related Disorders. 2023;110:105364.

[39] Priori A, Foffani G, Pesenti A, Tamma F, Bianchi AM, Pellegrini M, et al. Rhythm-specific pharmacological modulation of subthalamic activity in Parkinson’s disease. Exp Neurol. 2004;189(2):369–79.

[40] Neumann WJ, Degen K, Schneider GH, Brücke C, Huebl J, Brown P, et al. Subthalamic synchronized oscillatory activity correlates with motor impairment in patients with Parkinson’s disease. Mov Disord. 2016;31(11):1748–51.

[41] Pozzi NG, Isaias IU. Adaptive deep brain stimulation: Retuning Parkinson’s disease. Handb Clin Neurol. 2022;184:273–84.

[42] Okun MS, Gallo BV, Mandybur G, Jagid J, Foote KD, Revilla FJ, et al. Subthalamic deep brain stimulation with a constant-current device in Parkinson’s disease: an open-label randomised controlled trial. Lancet Neurol. 2012;11(2):140–9.

[43] Muller M, Scafa S, Hanafi I, Varescon C, Palmisano C, van der Gaag S, et al. Online prediction of optimal deep brain stimulation contacts from local field potentials in chronically-implanted w P k ’ Rx v. 2024:2024.11.26.24317968.

