## Supplementary file 1 for "From subthalamic local field potentials to the selection of chronic deep brain stimulation contacts in Parkinson’s disease - A systematic review"

### Supplementary methods

<u>Search strings used across the respective databases:</u>

**PubMed**
(("Parkinson Disease"[Mesh] OR "Parkinson*"[tw] **OR "Parkinsonian Disorders"[Mesh] OR "parkinsonian"[tw] OR "parkinsonism"[tw] OR "parkinson disease"[tw] OR "parkinson's disease"[tw] OR "parkinsons disease"[tw] OR "Paralysis Agitans"[tw] OR "parkinson"[tw] OR "parkinson*"[tw])** AND ("local field potential"[tw] OR "local field potentials"[tw] OR "local field potential*"[tw] OR ("local field"[tw] AND ("potential*"[tw] OR "Evoked Potentials"[mesh] OR "Membrane Potentials"[mesh])) OR "LFP"[tw] OR "feedback control"[tw] OR "feedback controls"[tw] OR "feedback controlled"[tw] OR "feedback control*"[tw] OR "neuronal activity"[tw] OR "neuronal activ*"[tw] OR "oscillation"[tw] OR "oscillations"[tw] OR "oscillation*"[tw] OR "oscillate"[tw] OR "oscillated"[tw] OR "oscillat*"[tw] OR "local field potential"[title/abstract:~4] OR "local field potentials"[title/abstract:~4] OR "feedback control"[title/abstract:~3] OR "feedback controls"[title/abstract:~3] OR "feedback controlled"[title/abstract:~3] OR "neuronal activity"[title/abstract:~3]) AND **("Deep Brain Stimulation"[Mesh] OR "Deep Brain Stimulation"[tw] OR "Deep Brain Stimulations"[tw] OR "Deep Brain Stimulator"[tw] OR "Deep Brain Stimulator"[tw] OR "DBS"[tw] OR "Deep Brain Stimulat*"[tw] OR "Deep Brain Stimulation"[title/abstract:~4] OR "Deep Brain Stimulations"[title/abstract:~4] OR "Deep Brain Stimulator"[title/abstract:~4] OR "Deep Brain Stimulator"[title/abstract:~4])** AND ("DBS program*"[tw] OR "program"[tw] OR "programs"[tw] OR "programme"[tw] OR "programmes"[tw] OR "programming"[tw] OR "program*"[tw] OR "optimization"[tw] OR "optimizing"[tw] OR "optimize"[tw] OR "optimiz*"[tw] OR "optimisation"[tw] OR "optimising"[tw] OR "optimising"[tw] OR "optimise"[tw] OR "optimis*"[tw] OR "contact point"[tw] OR "contact points"[tw] OR "chosen electrode"[tw] OR "chosen electrodes"[tw] OR "chosen electrode*"[tw] OR "monopolar review"[tw] OR "monopolar reviews"[tw] OR "monopolar review*"[tw] OR "chronic stimulation"[tw] OR "chronic stimulations"[tw] OR "chronic stimulat*"[tw] OR "stimulation parameter"[tw] OR "stimulation parameters"[tw] OR "stimulation parameter*"[tw] OR "contact point"[title/abstract:~3] OR "contact points"[title/abstract:~3] OR "chosen electrode"[title/abstract:~3] OR "chosen electrodes"[title/abstract:~3] OR "monopolar review"[title/abstract:~3] OR "monopolar reviews"[title/abstract:~3] OR "chronic stimulation"[title/abstract:~3] OR "chronic stimulations"[title/abstract:~3] OR "stimulation parameter"[title/abstract:~3] OR "stimulation parameters"[title/abstract:~3] **OR "contact selection"[tw] OR "contact choice"[title/abstract:~3] OR "contact selection"[title/abstract:~3] OR (("Electrodes, Implanted"[mesh] OR "Electrodes"[mesh] OR "Electrode"[tw] OR "Electrodes"[tw] OR "Macroelectrode"[tw] OR "Macroelectrodes"[tw]) AND ("contact"[tw] OR "contacts"[tw])) OR "localization"[tw] OR "localizing"[tw] OR "localized"[tw] OR "localize"[tw] OR "localiz*"[tw] OR "localisation"[tw] OR "localising"[tw] OR "localised"[tw] OR "localise"[tw] OR "localis*"[tw]**) NOT (("Animals"[mesh] OR **"veterinary"[ti] OR "rabbit"[ti] OR "rabbits"[ti] OR "animal"[ti] OR "animals"[ti] OR "mouse"[ti] OR "mice"[ti] OR "rodent"[ti] OR "rodents"[ti] OR "rat"[ti] OR "rats"[ti] OR "pig"[ti] OR "pigs"[ti] OR "porcine"[ti] OR "horse"[ti] OR "horses"[ti] OR "equine"[ti] OR "cow"[ti]**

**OR "cows"[ti] OR "bovine"[ti] OR "goat"[ti] OR "goats"[ti] OR "sheep"[ti] OR "ovine"[ti] OR "canine"[ti] OR "dog"[ti] OR "dogs"[ti] OR "feline"[ti] OR "cat"[ti] OR "cats"[ti])** NOT "Humans"[mesh]) AND english[la])

**Embase**
((exp *"Parkinson Disease"/ OR "Parkinson*".ti,ab **OR exp *"Parkinsonism"/ OR "parkinsonian".ti,ab OR "parkinsonism".ti,ab OR "parkinson disease".ti,ab OR "parkinson's disease".ti,ab OR "parkinsons disease".ti,ab OR "Paralysis Agitans".ti,ab OR "parkinson".ti,ab OR "parkinson*".ti,ab**) AND (*"local field potential"/ OR "local field potential".ti,ab OR "local field potentials".ti,ab OR "local field potential*".ti,ab OR "LFP".ti,ab OR "feedback control".ti,ab OR "feedback controls".ti,ab OR "feedback controlled".ti,ab OR "feedback control*".ti,ab OR "neuronal activity".ti,ab OR "neuronal activ*".ti,ab OR *"oscillation"/ OR "oscillation".ti,ab OR "oscillations".ti,ab OR "oscillation*".ti,ab OR "oscillate".ti,ab OR "oscillated".ti,ab OR "oscillat*".ti,ab OR (("local" ADJ4 "field" ADJ4 "potential") OR ("local" ADJ4 "field" ADJ4 "potentials") OR ("feedback" ADJ3 "control") OR ("feedback" ADJ3 "controls") OR ("feedback" ADJ3 "controlled") OR ("neuronal" ADJ3 "activity")).ti,ab) AND **(*"brain depth stimulation"/ OR "Deep Brain Stimulation".ti,ab OR "Deep Brain Stimulations".ti,ab OR "Deep Brain Stimulator".ti,ab OR "Deep Brain Stimulator".ti,ab OR "DBS".ti,ab OR "Deep Brain Stimulat*".ti,ab OR (("Deep" ADJ3 "Brain" ADJ3 "Stimulation") OR ("Deep" ADJ3 "Brain" ADJ3 "Stimulations") OR ("Deep" ADJ3 "Brain" ADJ3 "Stimulator") OR ("Deep" ADJ3 "Brain" ADJ3 "Stimulator")).ti,ab)** AND ("DBS program*".ti,ab OR "program".ti,ab OR "programs".ti,ab OR "programme".ti,ab OR "programmes".ti,ab OR "programming".ti,ab OR "program*".ti,ab OR "optimization".ti,ab OR "optimizing".ti,ab OR "optimize".ti,ab OR "optimiz*".ti,ab OR "optimisation".ti,ab OR "optimising".ti,ab OR "optimising".ti,ab OR "optimise".ti,ab OR "optimis*".ti,ab OR "contact point".ti,ab OR "contact points".ti,ab OR "chosen electrode".ti,ab OR "chosen electrodes".ti,ab OR "chosen electrode*".ti,ab OR "monopolar review".ti,ab OR "monopolar reviews".ti,ab OR "monopolar review*".ti,ab OR "chronic stimulation".ti,ab OR "chronic stimulations".ti,ab OR "chronic stimulat*".ti,ab OR "stimulation parameter".ti,ab OR "stimulation parameters".ti,ab OR "stimulation parameter*".ti,ab OR **"contact selection".ti,ab** OR (("contact" ADJ3 "point") OR ("contact" ADJ3 "points") OR ("chosen" ADJ3 "electrode") OR ("chosen" ADJ3 "electrodes") OR ("monopolar" ADJ3 "review") OR ("monopolar" ADJ3 "reviews") OR ("chronic" ADJ3 "stimulation") OR ("chronic" ADJ3 "stimulations") OR ("stimulation" ADJ3 "parameter") OR ("stimulation" ADJ3 "parameters") **OR ("contact" ADJ3 "choice") OR ("contact" ADJ3 "selection")).ti,ab OR "localization".ti,ab OR "localizing".ti,ab OR "localized".ti,ab OR "localize".ti,ab OR "localiz*".ti,ab OR "localisation".ti,ab OR "localising".ti,ab OR "localised".ti,ab OR "localise".ti,ab OR "localis*".ti,ab**) NOT ((exp "Animals"/ OR **"veterinary".ti OR "rabbit".ti OR "rabbits".ti OR "animal".ti OR "animals".ti OR "mouse".ti OR "mice".ti OR "rodent".ti OR "rodents".ti OR "rat".ti OR "rats".ti OR "pig".ti OR "pigs".ti OR "porcine".ti OR "horse".ti OR "horses".ti OR "equine".ti OR "cow".ti OR "cows".ti OR "bovine".ti OR "goat".ti OR "goats".ti OR "sheep".ti OR "ovine".ti OR "canine".ti OR "dog".ti OR "dogs".ti OR "feline".ti OR "cat".ti OR "cats".ti)** NOT exp "Humans"/) AND english.la)

**Web of Science**
((TI=("Parkinson Disease" OR "Parkinson*" **OR "Parkinsonism" OR "parkinsonian" OR "parkinsonism" OR "parkinson disease" OR "parkinson's disease" OR "parkinsons disease" OR "Paralysis Agitans" OR "parkinson" OR "parkinson*"**) OR AK=("Parkinson Disease" OR "Parkinson*" **OR "Parkinsonism" OR "parkinsonian" OR "parkinsonism"**

**OR "parkinson disease" OR "parkinson's disease" OR "parkinsons disease" OR "Paralysis Agitans" OR "parkinson" OR "parkinson*"**) OR AB=("Parkinson Disease" OR "Parkinson*" **OR "Parkinsonism" OR "parkinsonian" OR "parkinsonism" OR "parkinson disease" OR "parkinson's disease" OR "parkinsons disease" OR "Paralysis Agitans"**)) AND (TI=("local field potential" OR "local field potential" OR "local field potentials" OR "local field potential*" OR "LFP" OR "feedback control" OR "feedback controls" OR "feedback controlled" OR "feedback control*" OR "neuronal activity" OR "neuronal activ*" OR "oscillation" OR "oscillation" OR "oscillations" OR "oscillation*" OR "oscillate" OR "oscillated" OR "oscillat*" OR (("local" NEAR/4 "field" NEAR/4 "potential") OR ("local" NEAR/4 "field" NEAR/4 "potentials") OR ("feedback" NEAR/3 "control") OR ("feedback" NEAR/3 "controls") OR ("feedback" NEAR/3 "controlled") OR ("neuronal" NEAR/3 "activity"))) OR AK=("local field potential" OR "local field potential" OR "local field potentials" OR "local field potential*" OR "LFP" OR "feedback control" OR "feedback controls" OR "feedback controlled" OR "feedback control*" OR "neuronal activity" OR "neuronal activ*" OR "oscillation" OR "oscillation" OR "oscillations" OR "oscillation*" OR "oscillate" OR "oscillated" OR "oscillat*" OR (("local" NEAR/4 "field" NEAR/4 "potential") OR ("local" NEAR/4 "field" NEAR/4 "potentials") OR ("feedback" NEAR/3 "control") OR ("feedback" NEAR/3 "controls") OR ("feedback" NEAR/3 "controlled") OR ("neuronal" NEAR/3 "activity"))) OR AB=("local field potential" OR "local field potential" OR "local field potentials" OR "local field potential*" OR "LFP" OR "feedback control" OR "feedback controls" OR "feedback controlled" OR "feedback control*" OR "neuronal activity" OR "neuronal activ*" OR "oscillation" OR "oscillation" OR "oscillations" OR "oscillation*" OR "oscillate" OR "oscillated" OR "oscillat*" OR (("local" NEAR/4 "field" NEAR/4 "potential") OR ("local" NEAR/4 "field" NEAR/4 "potentials") OR ("feedback" NEAR/3 "control") OR ("feedback" NEAR/3 "controls") OR ("feedback" NEAR/3 "controlled") OR ("neuronal" NEAR/3 "activity")))) AND (TI=**("brain depth stimulation" OR "Deep Brain Stimulation" OR "Deep Brain Stimulations" OR "Deep Brain Stimulator" OR "Deep Brain Stimulator" OR "DBS" OR "Deep Brain Stimulat*" OR (("Deep" NEAR/3 "Brain" NEAR/3 "Stimulation") OR ("Deep" NEAR/3 "Brain" NEAR/3 "Stimulations") OR ("Deep" NEAR/3 "Brain" NEAR/3 "Stimulator") OR ("Deep" NEAR/3 "Brain" NEAR/3 "Stimulator")))** OR AK=**("brain depth stimulation" OR "Deep Brain Stimulation" OR "Deep Brain Stimulations" OR "Deep Brain Stimulator" OR "Deep Brain Stimulator" OR "DBS" OR "Deep Brain Stimulat*" OR (("Deep" NEAR/3 "Brain" NEAR/3 "Stimulation") OR ("Deep" NEAR/3 "Brain" NEAR/3 "Stimulations") OR ("Deep" NEAR/3 "Brain" NEAR/3 "Stimulator") OR ("Deep" NEAR/3 "Brain" NEAR/3 "Stimulator")))** OR AB=**("brain depth stimulation" OR "Deep Brain Stimulation" OR "Deep Brain Stimulations" OR "Deep Brain Stimulator" OR "Deep Brain Stimulator" OR "DBS" OR "Deep Brain Stimulat*" OR (("Deep" NEAR/3 "Brain" NEAR/3 "Stimulation") OR ("Deep" NEAR/3 "Brain" NEAR/3 "Stimulations") OR ("Deep" NEAR/3 "Brain" NEAR/3 "Stimulator") OR ("Deep" NEAR/3 "Brain" NEAR/3 "Stimulator"))))** AND (TI=("DBS program*" OR "program" OR "programs" OR "programme" OR "programmes" OR "programming" OR "program*" OR "optimization" OR "optimizing" OR "optimize" OR "optimiz*" OR "optimisation" OR "optimising" OR "optimising" OR "optimise" OR "optimis*" OR "contact point" OR "contact points" OR "chosen electrode" OR "chosen electrodes" OR "chosen electrode*" OR "monopolar review" OR "monopolar reviews" OR "monopolar review*" OR "chronic stimulation" OR "chronic stimulations" OR "chronic stimulat*" OR "stimulation parameter" OR "stimulation parameters" OR "stimulation parameter*" OR **"contact selection"** OR (("contact" NEAR/3 "point") OR ("contact" NEAR/3 "points") OR ("chosen" NEAR/3 "electrode") OR ("chosen" NEAR/3 "electrodes") OR ("monopolar" NEAR/3 "review") OR ("monopolar" NEAR/3 "reviews") OR ("chronic" NEAR/3 "stimulation") OR ("chronic" NEAR/3 "stimulations") OR ("stimulation" NEAR/3 "parameter")

OR ("stimulation" NEAR/3 "parameters"**) OR ("contact" NEAR/3 "choice") OR ("contact" NEAR/3 "selection")) OR "localization" OR "localizing" OR "localized" OR "localize" OR "localiz*" OR "localisation" OR "localising" OR "localised" OR "localise" OR "localis*"**) OR AK=("DBS program*" OR "program" OR "programs" OR "programme" OR "programmes" OR "programming" OR "program*" OR "optimization" OR "optimizing" OR "optimize" OR "optimiz*" OR "optimisation" OR "optimising" OR "optimising" OR "optimise" OR "optimis*" OR "contact point" OR "contact points" OR "chosen electrode" OR "chosen electrodes" OR "chosen electrode*" OR "monopolar review" OR "monopolar reviews" OR "monopolar review*" OR "chronic stimulation" OR "chronic stimulations" OR "chronic stimulat*" OR "stimulation parameter" OR "stimulation parameters" OR "stimulation parameter*" OR **"contact selection"** OR (("contact" NEAR/3 "point") OR ("contact" NEAR/3 "points") OR ("chosen" NEAR/3 "electrode") OR ("chosen" NEAR/3 "electrodes") OR ("monopolar" NEAR/3 "review") OR ("monopolar" NEAR/3 "reviews") OR ("chronic" NEAR/3 "stimulation") OR ("chronic" NEAR/3 "stimulations") OR ("stimulation" NEAR/3 "parameter") OR ("stimulation" NEAR/3 "parameters"**) OR ("contact" NEAR/3 "choice") OR ("contact" NEAR/3 "selection")) OR "localization" OR "localizing" OR "localized" OR "localize" OR "localiz*" OR "localisation" OR "localising" OR "localised" OR "localise" OR "localis*"**) OR AB=("DBS program*" OR "program" OR "programs" OR "programme" OR "programmes" OR "programming" OR "program*" OR "optimization" OR "optimizing" OR "optimize" OR "optimiz*" OR "optimisation" OR "optimising" OR "optimising" OR "optimise" OR "optimis*" OR "contact point" OR "contact points" OR "chosen electrode" OR "chosen electrodes" OR "chosen electrode*" OR "monopolar review" OR "monopolar reviews" OR "monopolar review*" OR "chronic stimulation" OR "chronic stimulations" OR "chronic stimulat*" OR "stimulation parameter" OR "stimulation parameters" OR "stimulation parameter*" OR **"contact selection"** OR (("contact" NEAR/3 "point") OR ("contact" NEAR/3 "points") OR ("chosen" NEAR/3 "electrode") OR ("chosen" NEAR/3 "electrodes") OR ("monopolar" NEAR/3 "review") OR ("monopolar" NEAR/3 "reviews") OR ("chronic" NEAR/3 "stimulation") OR ("chronic" NEAR/3 "stimulations") OR ("stimulation" NEAR/3 "parameter") OR ("stimulation" NEAR/3 "parameters"**) OR ("contact" NEAR/3 "choice") OR ("contact" NEAR/3 "selection")) OR "localization" OR "localizing" OR "localized" OR "localize" OR "localiz*" OR "localisation" OR "localising" OR "localised" OR "localise" OR "localis*"**)) NOT TI=(**"veterinary" OR "rabbit" OR "rabbits" OR "animal" OR "animals" OR "mouse" OR "mice" OR "rodent" OR "rodents" OR "rat" OR "rats" OR "pig" OR "pigs" OR "porcine" OR "horse" OR "horses" OR "equine" OR "cow" OR "cows" OR "bovine" OR "goat" OR "goats" OR "sheep" OR "ovine" OR "canine" OR "dog" OR "dogs" OR "feline" OR "cat" OR "cats"**) AND LA=english NOT DT=meeting abstract)

**Cochrane**

(("Parkinson Disease" OR "Parkinson" OR "Parkinson*" **OR "Parkinsonism" OR "parkinsonian" OR "parkinsonism" OR "parkinson disease" OR "parkinson's disease" OR "parkinsons disease" OR "Paralysis Agitans"**) AND ("local field potential" OR "local field potential" OR "local field potentials" OR "local field potential*" OR "LFP" OR "feedback control" OR "feedback controls" OR "feedback controlled" OR "feedback control*" OR "neuronal activity" OR "neuronal activ*" OR "oscillation" OR "oscillation" OR "oscillations" OR "oscillation*" OR "oscillate" OR "oscillated" OR "oscillat*" OR (("local" NEAR/4 "field" NEAR/4 "potential") OR ("local" NEAR/4 "field" NEAR/4 "potentials") OR ("feedback" NEAR/3 "control") OR ("feedback" NEAR/3 "controls") OR ("feedback" NEAR/3 "controlled") OR ("neuronal" NEAR/3 "activity"))) AND (**"brain depth stimulation" OR "Deep Brain Stimulation" OR "Deep Brain Stimulations" OR "Deep Brain Stimulator" OR "Deep Brain Stimulator" OR "DBS" OR "Deep Brain Stimulat*" OR (("Deep" NEAR/3 "Brain"**

**NEAR/3 "Stimulation") OR ("Deep" NEAR/3 "Brain" NEAR/3 "Stimulations") OR ("Deep" NEAR/3 "Brain" NEAR/3 "Stimulator") OR ("Deep" NEAR/3 "Brain" NEAR/3 "Stimulator")))** AND ("DBS program*" OR "program" OR "programs" OR "programme" OR "programmes" OR "programming" OR "program*" OR "optimization" OR "optimizing" OR "optimize" OR "optimiz*" OR "optimisation" OR "optimising" OR "optimising" OR "optimise" OR "optimis*" OR "contact point" OR "contact points" OR "chosen electrode" OR "chosen electrodes" OR "chosen electrode*" OR "monopolar review" OR "monopolar reviews" OR "monopolar review*" OR "chronic stimulation" OR "chronic stimulations" OR "chronic stimulat*" OR "stimulation parameter" OR "stimulation parameters" OR "stimulation parameter*" OR **"contact selection"** OR (("contact" NEAR/3 "point") OR ("contact" NEAR/3 "points") OR ("chosen" NEAR/3 "electrode") OR ("chosen" NEAR/3 "electrodes") OR ("monopolar" NEAR/3 "review") OR ("monopolar" NEAR/3 "reviews") OR ("chronic" NEAR/3 "stimulation") OR ("chronic" NEAR/3 "stimulations") OR ("stimulation" NEAR/3 "parameter") OR ("stimulation" NEAR/3 "parameters"**) OR ("contact" NEAR/3 "choice") OR ("contact" NEAR/3 "selection")) OR "localization" OR "localizing" OR "localized" OR "localize" OR "localiz*" OR "localisation" OR "localising" OR "localised" OR "localise" OR "localis*"**)):ti,ab,kw)

AND LA=english

**Emcare**
((exp *"Parkinson Disease"/ OR "Parkinson*".ti,ab **OR exp *"Parkinsonism"/ OR "parkinsonian".ti,ab OR "parkinsonism".ti,ab OR "parkinson disease".ti,ab OR "parkinson's disease".ti,ab OR "parkinsons disease".ti,ab OR "Paralysis Agitans".ti,ab OR "parkinson".ti,ab OR "parkinson*".ti,ab**) AND (*"local field potential"/ OR "local field potential".ti,ab OR "local field potentials".ti,ab OR "local field potential*".ti,ab OR "LFP".ti,ab OR "feedback control".ti,ab OR "feedback controls".ti,ab OR "feedback controlled".ti,ab OR "feedback control*".ti,ab OR "neuronal activity".ti,ab OR "neuronal activ*".ti,ab OR *"oscillation"/ OR "oscillation".ti,ab OR "oscillations".ti,ab OR "oscillation*".ti,ab OR "oscillate".ti,ab OR "oscillated".ti,ab OR "oscillat*".ti,ab OR (("local" ADJ4 "field" ADJ4 "potential") OR ("local" ADJ4 "field" ADJ4 "potentials") OR ("feedback" ADJ3 "control") OR ("feedback" ADJ3 "controls") OR ("feedback" ADJ3 "controlled") OR ("neuronal" ADJ3 "activity")).ti,ab) AND **(*"brain depth stimulation"/ OR "Deep Brain Stimulation".ti,ab OR "Deep Brain Stimulations".ti,ab OR "Deep Brain Stimulator".ti,ab OR "Deep Brain Stimulator".ti,ab OR "DBS".ti,ab OR "Deep Brain Stimulat*".ti,ab OR (("Deep" ADJ3 "Brain" ADJ3 "Stimulation") OR ("Deep" ADJ3 "Brain" ADJ3 "Stimulations") OR ("Deep" ADJ3 "Brain" ADJ3 "Stimulator") OR ("Deep" ADJ3 "Brain" ADJ3 "Stimulator")).ti,ab)** AND ("DBS program*".ti,ab OR "program".ti,ab OR "programs".ti,ab OR "programme".ti,ab OR "programmes".ti,ab OR "programming".ti,ab OR "program*".ti,ab OR "optimization".ti,ab OR "optimizing".ti,ab OR "optimize".ti,ab OR "optimiz*".ti,ab OR "optimisation".ti,ab OR "optimising".ti,ab OR "optimising".ti,ab OR "optimise".ti,ab OR "optimis*".ti,ab OR "contact point".ti,ab OR "contact points".ti,ab OR "chosen electrode".ti,ab OR "chosen electrodes".ti,ab OR "chosen electrode*".ti,ab OR "monopolar review".ti,ab OR "monopolar reviews".ti,ab OR "monopolar review*".ti,ab OR "chronic stimulation".ti,ab OR "chronic stimulations".ti,ab OR "chronic stimulat*".ti,ab OR "stimulation parameter".ti,ab OR "stimulation parameters".ti,ab OR "stimulation parameter*".ti,ab OR **"contact selection".ti,ab** OR (("contact" ADJ3 "point") OR ("contact" ADJ3 "points") OR ("chosen" ADJ3 "electrode") OR ("chosen" ADJ3 "electrodes") OR ("monopolar" ADJ3 "review") OR ("monopolar" ADJ3 "reviews") OR ("chronic" ADJ3 "stimulation") OR ("chronic" ADJ3 "stimulations") OR ("stimulation" ADJ3 "parameter") OR ("stimulation" ADJ3 "parameters"**) OR ("contact" ADJ3 "choice") OR ("contact" ADJ3**

**"selection")).ti,ab OR "localization".ti,ab OR "localizing".ti,ab OR "localized".ti,ab OR "localize".ti,ab OR "localiz*".ti,ab OR "localisation".ti,ab OR "localising".ti,ab OR "localised".ti,ab OR "localise".ti,ab OR "localis*".ti,ab**) NOT ((exp "Animals"/ OR **"veterinary".ti OR "rabbit".ti OR "rabbits".ti OR "animal".ti OR "animals".ti OR "mouse".ti OR "mice".ti OR "rodent".ti OR "rodents".ti OR "rat".ti OR "rats".ti OR "pig".ti OR "pigs".ti OR "porcine".ti OR "horse".ti OR "horses".ti OR "equine".ti OR "cow".ti OR "cows".ti OR "bovine".ti OR "goat".ti OR "goats".ti OR "sheep".ti OR "ovine".ti OR "canine".ti OR "dog".ti OR "dogs".ti OR "feline".ti OR "cat".ti OR "cats".ti)** NOT exp "Humans"/) AND english.la)

**PsycINFO**

((TI("Parkinson Disease" OR "Parkinson*" **OR "Parkinsonism" OR "parkinsonian" OR "parkinsonism" OR "parkinson disease" OR "parkinson's disease" OR "parkinsons disease" OR "Paralysis Agitans" OR "parkinson" OR "parkinson*"**) OR SU("Parkinson Disease" OR "Parkinson*" **OR "Parkinsonism" OR "parkinsonian" OR "parkinsonism" OR "parkinson disease" OR "parkinson's disease" OR "parkinsons disease" OR "Paralysis Agitans" OR "parkinson" OR "parkinson*"**) OR MA("Parkinson Disease" OR "Parkinson*" **OR "Parkinsonism" OR "parkinsonian" OR "parkinsonism" OR "parkinson disease" OR "parkinson's disease" OR "parkinsons disease" OR "Paralysis Agitans" OR "parkinson" OR "parkinson*"**) OR AB("Parkinson Disease" OR "Parkinson*" **OR "Parkinsonism" OR "parkinsonian" OR "parkinsonism" OR "parkinson disease" OR "parkinson's disease" OR "parkinsons disease" OR "Paralysis Agitans"**)) AND (TI("local field potential" OR "local field potential" OR "local field potentials" OR "local field potential*" OR "LFP" OR "feedback control" OR "feedback controls" OR "feedback controlled" OR "feedback control*" OR "neuronal activity" OR "neuronal activ*" OR "oscillation" OR "oscillation" OR "oscillations" OR "oscillation*" OR "oscillate" OR "oscillated" OR "oscillat*" OR (("local" N4 "field" N4 "potential") OR ("local" N4 "field" N4 "potentials") OR ("feedback" N3 "control") OR ("feedback" N3 "controls") OR ("feedback" N3 "controlled") OR ("neuronal" N3 "activity"))) OR SU("local field potential" OR "local field potential" OR "local field potentials" OR "local field potential*" OR "LFP" OR "feedback control" OR "feedback controls" OR "feedback controlled" OR "feedback control*" OR "neuronal activity" OR "neuronal activ*" OR "oscillation" OR "oscillation" OR "oscillations" OR "oscillation*" OR "oscillate" OR "oscillated" OR "oscillat*" OR (("local" N4 "field" N4 "potential") OR ("local" N4 "field" N4 "potentials") OR ("feedback" N3 "control") OR ("feedback" N3 "controls") OR ("feedback" N3 "controlled") OR ("neuronal" N3 "activity"))) OR MA("local field potential" OR "local field potential" OR "local field potentials" OR "local field potential*" OR "LFP" OR "feedback control" OR "feedback controls" OR "feedback controlled" OR "feedback control*" OR "neuronal activity" OR "neuronal activ*" OR "oscillation" OR "oscillation" OR "oscillations" OR "oscillation*" OR "oscillate" OR "oscillated" OR "oscillat*" OR (("local" N4 "field" N4 "potential") OR ("local" N4 "field" N4 "potentials") OR ("feedback" N3 "control") OR ("feedback" N3 "controls") OR ("feedback" N3 "controlled") OR ("neuronal" N3 "activity"))) OR AB("local field potential" OR "local field potential" OR "local field potentials" OR "local field potential*" OR "LFP" OR "feedback control" OR "feedback controls" OR "feedback controlled" OR "feedback control*" OR "neuronal activity" OR "neuronal activ*" OR "oscillation" OR "oscillation" OR "oscillations" OR "oscillation*" OR "oscillate" OR "oscillated" OR "oscillat*" OR (("local" N4 "field" N4 "potential") OR ("local" N4 "field" N4 "potentials") OR ("feedback" N3 "control") OR ("feedback" N3 "controls") OR ("feedback" N3 "controlled") OR ("neuronal" N3 "activity")))) AND (TI**("brain depth stimulation" OR "Deep Brain Stimulation" OR "Deep Brain Stimulations" OR "Deep Brain Stimulator" OR "Deep Brain Stimulator" OR "DBS" OR "Deep Brain Stimulat*"**

**OR (("Deep" N3 "Brain" N3 "Stimulation") OR ("Deep" N3 "Brain" N3 "Stimulations") OR ("Deep" N3 "Brain" N3 "Stimulator") OR ("Deep" N3 "Brain" N3 "Stimulator")))** OR SU**("brain depth stimulation" OR "Deep Brain Stimulation" OR "Deep Brain Stimulations" OR "Deep Brain Stimulator" OR "Deep Brain Stimulator" OR "DBS" OR "Deep Brain Stimulat*" OR (("Deep" N3 "Brain" N3 "Stimulation") OR ("Deep" N3 "Brain" N3 "Stimulations") OR ("Deep" N3 "Brain" N3 "Stimulator") OR ("Deep" N3 "Brain" N3 "Stimulator")))** OR MA**("brain depth stimulation" OR "Deep Brain Stimulation" OR "Deep Brain Stimulations" OR "Deep Brain Stimulator" OR "Deep Brain Stimulator" OR "DBS" OR "Deep Brain Stimulat*" OR (("Deep" N3 "Brain" N3 "Stimulation") OR ("Deep" N3 "Brain" N3 "Stimulations") OR ("Deep" N3 "Brain" N3 "Stimulator") OR ("Deep" N3 "Brain" N3 "Stimulator")))** OR AB**("brain depth stimulation" OR "Deep Brain Stimulation" OR "Deep Brain Stimulations" OR "Deep Brain Stimulator" OR "Deep Brain Stimulator" OR "DBS" OR "Deep Brain Stimulat*" OR (("Deep" N3 "Brain" N3 "Stimulation") OR ("Deep" N3 "Brain" N3 "Stimulations") OR ("Deep" N3 "Brain" N3 "Stimulator") OR ("Deep" N3 "Brain" N3 "Stimulator"))))** AND (TI("DBS program*" OR "program" OR "programs" OR "programme" OR "programmes" OR "programming" OR "program*" OR "optimization" OR "optimizing" OR "optimize" OR "optimiz*" OR "optimisation" OR "optimising" OR "optimising" OR "optimise" OR "optimis*" OR "contact point" OR "contact points" OR "chosen electrode" OR "chosen electrodes" OR "chosen electrode*" OR "monopolar review" OR "monopolar reviews" OR "monopolar review*" OR "chronic stimulation" OR "chronic stimulations" OR "chronic stimulat*" OR "stimulation parameter" OR "stimulation parameters" OR "stimulation parameter*" OR **"contact selection"** OR (("contact" N3 "point") OR ("contact" N3 "points") OR ("chosen" N3 "electrode") OR ("chosen" N3 "electrodes") OR ("monopolar" N3 "review") OR ("monopolar" N3 "reviews") OR ("chronic" N3 "stimulation") OR ("chronic" N3 "stimulations") OR ("stimulation" N3 "parameter") OR ("stimulation" N3 "parameters"**) OR ("contact" N3 "choice") OR ("contact" N3 "selection")) OR "localization" OR "localizing" OR "localized" OR "localize" OR "localiz*" OR "localisation" OR "localising" OR "localised" OR "localise" OR "localis*"**) OR SU("DBS program*" OR "program" OR "programs" OR "programme" OR "programmes" OR "programming" OR "program*" OR "optimization" OR "optimizing" OR "optimize" OR "optimiz*" OR "optimisation" OR "optimising" OR "optimising" OR "optimise" OR "optimis*" OR "contact point" OR "contact points" OR "chosen electrode" OR "chosen electrodes" OR "chosen electrode*" OR "monopolar review" OR "monopolar reviews" OR "monopolar review*" OR "chronic stimulation" OR "chronic stimulations" OR "chronic stimulat*" OR "stimulation parameter" OR "stimulation parameters" OR "stimulation parameter*" OR **"contact selection"** OR (("contact" N3 "point") OR ("contact" N3 "points") OR ("chosen" N3 "electrode") OR ("chosen" N3 "electrodes") OR ("monopolar" N3 "review") OR ("monopolar" N3 "reviews") OR ("chronic" N3 "stimulation") OR ("chronic" N3 "stimulations") OR ("stimulation" N3 "parameter") OR ("stimulation" N3 "parameters"**) OR ("contact" N3 "choice") OR ("contact" N3 "selection")) OR "localization" OR "localizing" OR "localized" OR "localize" OR "localiz*" OR "localisation" OR "localising" OR "localised" OR "localise" OR "localis*"**) OR MA("DBS program*" OR "program" OR "programs" OR "programme" OR "programmes" OR "programming" OR "program*" OR "optimization" OR "optimizing" OR "optimize" OR "optimiz*" OR "optimisation" OR "optimising" OR "optimising" OR "optimise" OR "optimis*" OR "contact point" OR "contact points" OR "chosen electrode" OR "chosen electrodes" OR "chosen electrode*" OR "monopolar review" OR "monopolar reviews" OR "monopolar review*" OR "chronic stimulation" OR "chronic stimulations" OR "chronic stimulat*" OR "stimulation parameter" OR "stimulation parameters" OR "stimulation parameter*" OR **"contact selection"** OR (("contact" N3 "point") OR ("contact" N3 "points") OR ("chosen" N3 "electrode") OR ("chosen" N3 "electrodes") OR ("monopolar" N3 "review")

OR ("monopolar" N3 "reviews") OR ("chronic" N3 "stimulation") OR ("chronic" N3 "stimulations") OR ("stimulation" N3 "parameter") OR ("stimulation" N3 "parameters"**) OR ("contact" N3 "choice") OR ("contact" N3 "selection")) OR "localization" OR "localizing" OR "localized" OR "localize" OR "localiz*" OR "localisation" OR "localising" OR "localised" OR "localise" OR "localis*"**) OR AB("DBS program*" OR "program" OR "programs" OR "programme" OR "programmes" OR "programming" OR "program*" OR "optimization" OR "optimizing" OR "optimize" OR "optimiz*" OR "optimisation" OR "optimising" OR "optimising" OR "optimise" OR "optimis*" OR "contact point" OR "contact points" OR "chosen electrode" OR "chosen electrodes" OR "chosen electrode*" OR "monopolar review" OR "monopolar reviews" OR "monopolar review*" OR "chronic stimulation" OR "chronic stimulations" OR "chronic stimulat*" OR "stimulation parameter" OR "stimulation parameters" OR "stimulation parameter*" OR **"contact selection"** OR (("contact" N3 "point") OR ("contact" N3 "points") OR ("chosen" N3 "electrode") OR ("chosen" N3 "electrodes") OR ("monopolar" N3 "review") OR ("monopolar" N3 "reviews") OR ("chronic" N3 "stimulation") OR ("chronic" N3 "stimulations") OR ("stimulation" N3 "parameter") OR ("stimulation" N3 "parameters"**) OR ("contact" N3 "choice") OR ("contact" N3 "selection")) OR "localization" OR "localizing" OR "localized" OR "localize" OR "localiz*" OR "localisation" OR "localising" OR "localised" OR "localise" OR "localis*"**)) NOT TI(**"veterinary" OR "rabbit" OR "rabbits" OR "animal" OR "animals" OR "mouse" OR "mice" OR "rodent" OR "rodents" OR "rat" OR "rats" OR "pig" OR "pigs" OR "porcine" OR "horse" OR "horses" OR "equine" OR "cow" OR "cows" OR "bovine" OR "goat" OR "goats" OR "sheep" OR "ovine" OR "canine" OR "dog" OR "dogs" OR "feline" OR "cat" OR "cats"**))

AND LA=english

**Academic Search Premier**

((TI("Parkinson Disease" OR "Parkinson*" **OR "Parkinsonism" OR "parkinsonian" OR "parkinsonism" OR "parkinson disease" OR "parkinson's disease" OR "parkinsons disease" OR "Paralysis Agitans" OR "parkinson" OR "parkinson*"**) OR SU("Parkinson Disease" OR "Parkinson*" **OR "Parkinsonism" OR "parkinsonian" OR "parkinsonism" OR "parkinson disease" OR "parkinson's disease" OR "parkinsons disease" OR "Paralysis Agitans" OR "parkinson" OR "parkinson*"**) OR KW("Parkinson Disease" OR "Parkinson*" **OR "Parkinsonism" OR "parkinsonian" OR "parkinsonism" OR "parkinson disease" OR "parkinson's disease" OR "parkinsons disease" OR "Paralysis Agitans" OR "parkinson" OR "parkinson*"**) OR AB("Parkinson Disease" OR "Parkinson*" **OR "Parkinsonism" OR "parkinsonian" OR "parkinsonism" OR "parkinson disease" OR "parkinson's disease" OR "parkinsons disease" OR "Paralysis Agitans"**)) AND (TI("local field potential" OR "local field potential" OR "local field potentials" OR "local field potential*" OR "LFP" OR "feedback control" OR "feedback controls" OR "feedback controlled" OR "feedback control*" OR "neuronal activity" OR "neuronal activ*" OR "oscillation" OR "oscillation" OR "oscillations" OR "oscillation*" OR "oscillate" OR "oscillated" OR "oscillat*" OR (("local" N4 "field" N4 "potential") OR ("local" N4 "field" N4 "potentials") OR ("feedback" N3 "control") OR ("feedback" N3 "controls") OR ("feedback" N3 "controlled") OR ("neuronal" N3 "activity"))) OR SU("local field potential" OR "local field potential" OR "local field potentials" OR "local field potential*" OR "LFP" OR "feedback control" OR "feedback controls" OR "feedback controlled" OR "feedback control*" OR "neuronal activity" OR "neuronal activ*" OR "oscillation" OR "oscillation" OR "oscillations" OR "oscillation*" OR "oscillate" OR "oscillated" OR "oscillat*" OR (("local" N4 "field" N4 "potential") OR ("local" N4 "field" N4 "potentials") OR ("feedback" N3 "control") OR ("feedback" N3 "controls") OR ("feedback" N3 "controlled") OR ("neuronal" N3 "activity")))

OR KW("local field potential" OR "local field potential" OR "local field potentials" OR "local field potential*" OR "LFP" OR "feedback control" OR "feedback controls" OR "feedback controlled" OR "feedback control*" OR "neuronal activity" OR "neuronal activ*" OR "oscillation" OR "oscillation" OR "oscillations" OR "oscillation*" OR "oscillate" OR "oscillated" OR "oscillat*" OR (("local" N4 "field" N4 "potential") OR ("local" N4 "field" N4 "potentials") OR ("feedback" N3 "control") OR ("feedback" N3 "controls") OR ("feedback" N3 "controlled") OR ("neuronal" N3 "activity"))) OR AB("local field potential" OR "local field potential" OR "local field potentials" OR "local field potential*" OR "LFP" OR "feedback control" OR "feedback controls" OR "feedback controlled" OR "feedback control*" OR "neuronal activity" OR "neuronal activ*" OR "oscillation" OR "oscillation" OR "oscillations" OR "oscillation*" OR "oscillate" OR "oscillated" OR "oscillat*" OR (("local" N4 "field" N4 "potential") OR ("local" N4 "field" N4 "potentials") OR ("feedback" N3 "control") OR ("feedback" N3 "controls") OR ("feedback" N3 "controlled") OR ("neuronal" N3 "activity")))) AND (TI**("brain depth stimulation" OR "Deep Brain Stimulation" OR "Deep Brain Stimulations" OR "Deep Brain Stimulator" OR "Deep Brain Stimulator" OR "DBS" OR "Deep Brain Stimulat*" OR (("Deep" N3 "Brain" N3 "Stimulation") OR ("Deep" N3 "Brain" N3 "Stimulations") OR ("Deep" N3 "Brain" N3 "Stimulator") OR ("Deep" N3 "Brain" N3 "Stimulator")))** OR SU**("brain depth stimulation" OR "Deep Brain Stimulation" OR "Deep Brain Stimulations" OR "Deep Brain Stimulator" OR "Deep Brain Stimulator" OR "DBS" OR "Deep Brain Stimulat*" OR (("Deep" N3 "Brain" N3 "Stimulation") OR ("Deep" N3 "Brain" N3 "Stimulations") OR ("Deep" N3 "Brain" N3 "Stimulator") OR ("Deep" N3 "Brain" N3 "Stimulator")))** OR KW**("brain depth stimulation" OR "Deep Brain Stimulation" OR "Deep Brain Stimulations" OR "Deep Brain Stimulator" OR "Deep Brain Stimulator" OR "DBS" OR "Deep Brain Stimulat*" OR (("Deep" N3 "Brain" N3 "Stimulation") OR ("Deep" N3 "Brain" N3 "Stimulations") OR ("Deep" N3 "Brain" N3 "Stimulator") OR ("Deep" N3 "Brain" N3 "Stimulator")))** OR AB**("brain depth stimulation" OR "Deep Brain Stimulation" OR "Deep Brain Stimulations" OR "Deep Brain Stimulator" OR "Deep Brain Stimulator" OR "DBS" OR "Deep Brain Stimulat*" OR (("Deep" N3 "Brain" N3 "Stimulation") OR ("Deep" N3 "Brain" N3 "Stimulations") OR ("Deep" N3 "Brain" N3 "Stimulator") OR ("Deep" N3 "Brain" N3 "Stimulator"))))** AND (TI("DBS program*" OR "program" OR "programs" OR "programme" OR "programmes" OR "programming" OR "program*" OR "optimization" OR "optimizing" OR "optimize" OR "optimiz*" OR "optimisation" OR "optimising" OR "optimising" OR "optimise" OR "optimis*" OR "contact point" OR "contact points" OR "chosen electrode" OR "chosen electrodes" OR "chosen electrode*" OR "monopolar review" OR "monopolar reviews" OR "monopolar review*" OR "chronic stimulation" OR "chronic stimulations" OR "chronic stimulat*" OR "stimulation parameter" OR "stimulation parameters" OR "stimulation parameter*" OR **"contact selection"** OR (("contact" N3 "point") OR ("contact" N3 "points") OR ("chosen" N3 "electrode") OR ("chosen" N3 "electrodes") OR ("monopolar" N3 "review") OR ("monopolar" N3 "reviews") OR ("chronic" N3 "stimulation") OR ("chronic" N3 "stimulations") OR ("stimulation" N3 "parameter") OR ("stimulation" N3 "parameters"**) OR ("contact" N3 "choice") OR ("contact" N3 "selection")) OR "localization" OR "localizing" OR "localized" OR "localize" OR "localiz*" OR "localisation" OR "localising" OR "localised" OR "localise" OR "localis*"**) OR SU("DBS program*" OR "program" OR "programs" OR "programme" OR "programmes" OR "programming" OR "program*" OR "optimization" OR "optimizing" OR "optimize" OR "optimiz*" OR "optimisation" OR "optimising" OR "optimising" OR "optimise" OR "optimis*" OR "contact point" OR "contact points" OR "chosen electrode" OR "chosen electrodes" OR "chosen electrode*" OR "monopolar review" OR "monopolar reviews" OR "monopolar review*" OR "chronic stimulation" OR "chronic stimulations" OR "chronic stimulat*" OR "stimulation parameter" OR "stimulation parameters" OR "stimulation parameter*" OR **"contact selection"** OR (("contact" N3 "point") OR ("contact" N3 "points") OR ("chosen" N3 "electrode") OR ("chosen" N3 "electrodes") OR ("monopolar" N3 "review") OR ("monopolar" N3 "reviews") OR ("chronic" N3 "stimulation") OR ("chronic" N3 "stimulations") OR ("stimulation" N3 "parameter") OR ("stimulation" N3 "parameters"**) OR ("contact" N3 "choice") OR ("contact" N3 "selection")) OR "localization" OR "localizing" OR "localized" OR "localize" OR "localiz*" OR "localisation" OR "localising" OR "localised" OR "localise" OR "localis*"**) OR KW("DBS program*" OR "program" OR "programs" OR "programme" OR "programmes" OR "programming" OR "program*" OR "optimization" OR "optimizing" OR "optimize" OR "optimiz*" OR "optimisation" OR "optimising" OR "optimising" OR "optimise" OR "optimis*" OR "contact point" OR "contact points" OR "chosen electrode" OR "chosen electrodes" OR "chosen electrode*" OR "monopolar review" OR "monopolar reviews" OR "monopolar review*" OR "chronic stimulation" OR "chronic stimulations" OR "chronic stimulat*" OR "stimulation parameter" OR "stimulation parameters" OR "stimulation parameter*" OR **"contact selection"** OR (("contact" N3 "point") OR ("contact" N3 "points") OR ("chosen" N3 "electrode") OR ("chosen" N3 "electrodes") OR ("monopolar" N3 "review") OR ("monopolar" N3 "reviews") OR ("chronic" N3 "stimulation") OR ("chronic" N3 "stimulations") OR ("stimulation" N3 "parameter") OR ("stimulation" N3 "parameters"**) OR ("contact" N3 "choice") OR ("contact" N3 "selection")) OR "localization" OR "localizing" OR "localized" OR "localize" OR "localiz*" OR "localisation" OR "localising" OR "localised" OR "localise" OR "localis*"**) OR AB("DBS program*" OR "program" OR "programs" OR "programme" OR "programmes" OR "programming" OR "program*" OR "optimization" OR "optimizing" OR "optimize" OR "optimiz*" OR "optimisation" OR "optimising" OR "optimising" OR "optimise" OR "optimis*" OR "contact point" OR "contact points" OR "chosen electrode" OR "chosen electrodes" OR "chosen electrode*" OR "monopolar review" OR "monopolar reviews" OR "monopolar review*" OR "chronic stimulation" OR "chronic stimulations" OR "chronic stimulat*" OR "stimulation parameter" OR "stimulation parameters" OR "stimulation parameter*" OR **"contact selection"** OR (("contact" N3 "point") OR ("contact" N3 "points") OR ("chosen" N3 "electrode") OR ("chosen" N3 "electrodes") OR ("monopolar" N3 "review") OR ("monopolar" N3 "reviews") OR ("chronic" N3 "stimulation") OR ("chronic" N3 "stimulations") OR ("stimulation" N3 "parameter") OR ("stimulation" N3 "parameters"**) OR ("contact" N3 "choice") OR ("contact" N3 "selection")) OR "localization" OR "localizing" OR "localized" OR "localize" OR "localiz*" OR "localisation" OR "localising" OR "localised" OR "localise" OR "localis*"**)) NOT TI(**"veterinary" OR "rabbit" OR "rabbits" OR "animal" OR "animals" OR "mouse" OR "mice" OR "rodent" OR "rodents" OR "rat" OR "rats" OR "pig" OR "pigs" OR "porcine" OR "horse" OR "horses" OR "equine" OR "cow" OR "cows" OR "bovine" OR "goat" OR "goats" OR "sheep" OR "ovine" OR "canine" OR "dog" OR "dogs" OR "feline" OR "cat" OR "cats"**))

AND LA=english

**Google Scholar**

- First 50 results
- Manual selection on human, language and meeting abstract

"Parkinson"|"Parkinsons" "local field potential"|"local field potentials"|"LFP"|"feedback control"|"neuronal activity"|"oscillation" **"Deep Brain Stimulation"** "contact" -animal|animals|rat|rats

### Supplementary results

#### Primary results

Two studies employed bipolar recordings and a bipolar *clinical stimulation contact* choice [1, 37], as they considered both the cathode and anode as a reference for the predictions, the *a priori* chance was, doubled. This was not applicable to the two other studies that also considered a bipolar *clinical stimulation contact* choice [26, 28], as they considered the chathode alone as reference for their predicitions.

Furthermore,  efforts were made to correctly distinguish between bipolar and monopolar recordings. However, misinterpretations may have occurred in articles where monopolar recordings were performed at various depths across the STN and/or were re-referenced offline to partially mimic bipolar recordings [1, 11, 13, 25].

#### Secundary results

<u>LFP-based programming vs. Empirical programming</u>

Five articles compared clinical results (UPDRS-III scores or clinical efficacy/symptom control) achieved by MPR-based DBS programming to clinical results obtained by beta-based contact choices [5, 27, 29, 30, 36]. Three of these articles, focussing on peak- or low-beta-activity, found no significant difference in clinical effect between the two programming entities [5, 27, 29, 30]. The research by J. Strelow et al. (2023) was the only study to individually compare high-beta based programming to empirical programming, here, this programming method achieved significantly lower clinical results than programming based on MPR [29]. in another study it was shown that the contact with the highest difference in beta/alpha ratio (when comparing stimulation ON and stimulation OFF recordings) achieved a significantly higher UPDRS-III improvement ratio compared to empirical programming [36]. Furthermore, LFP-based programming was found to require significantly less programming iterations than empirical programming [36]. Similar results were provided by T. Binder et al. (2023) [5]. Finally, the research conducted by F. Yoshida et al. (2010) demonstrates that using the clinically chosen chronic stimulation contacts requires higher stimulation voltages compared to contacts selected by means of beta-power evaluations (r=0.322, p=0.017) [11].

Effects of general anaesthesia

The effect of general anaesthesia (GA) on the performance of beta-band, evoked resonant neural activity (ERNA), and HFO-based contact predictions was evaluated by two studies. The research by N.C. Sinclair et al. (2021) revealed a decrease in predictive accuracy of beta-based and HFO-based predictions under GA compared to awake conditions (beta: awake 54.2% / GA 16.7%; HFO: awake 70.8% / GA 33.3%). This reduction was not present for ERNA-based predictions (awake 79.2% / GA 87.5%) which improved under GA [33]. The research by L. di Biase et al. (2023) did not reveal a negative effect of GA on the diagnostic performance of beta-power [23].

Movement vs. resting state

In five articles the effect of active or passive movement on the correlation between different frequencies from LFP recordings and the *clinical stimulation contact* were evaluated [1, 15, 21, 26, 34]. A single article evaluated the effect of passive movement and found the predictive accuracy of several aspects of the beta-band to be similar for both the resting and passive movement state [15]. Three articles investigated the effect of active movement (i.e. during specific hand movements) [1, 21, 26]. One of these studies found that resting (fast-) gamma, resting or moving-modulated beta, and movement-modulated HFO were most predictive for the *clinical stimulation contact* [21]. However, another article demonstrated that the position of the *clinical stimulation contact* is best predicted by the gamma-band during the movement state, whereas the predictive power of beta-band activity is greatest at rest [1]. This study also showed significant differences in beta-power between optimal and non-optimal *clinical stimulation contacts* in both movement and resting states (p<0.0001) [1]. In the research by, R. Kochanski et al (2019) no difference was observed in the performance of resting or movement state recordings [26]. However, this study did find an increase in beta-power during movement compared to rest in six out of ten hemispheres (p=0.05) [26].

The research by L. Busch et al. (2023) evaluated movement velocity in relation to stimulation-induced beta suppression and directional beta-power. They showed that movement velocity increases with stronger suppression of beta-power ($\beta = -0.008$, $t = -4.69$, $P < 0.001$) [34]. Moreover, higher directional beta-power with stimulation turned OFF was accompanied by stronger improvement in movement velocity when increasing stimulation amplitude ($\beta = 0.028$, $t = 2.28$, $P = 0.023$) [34]. Finally, a stronger correlation between movement velocity rankings and contact rankings based on stimulation-induced beta-suppression was found in comparison to directional beta-power (correlation difference: $F = 5.9$, $P = 0.019$) [34].

Non-frequency biomarkers

The performance of two non-frequency factors, anatomy and ERNA, was assessed across six articles. When considering anatomy, two articles, both from research group #2 report a predictive accuracy of 67% and 68% when ranking contacts based on imaging in order of proximity to a nominated ideal anatomical location [17, 20]. Furthermore, a study conducted by research group #1 showed that incorporating an anatomical landmark as additional feature could improve the predictive accuracy when considering the entire DBS lead [21]. This was, however, not the case when only considering the six segmented contacts. The research performed by L. di Biase et al. (2023) showed that combining beta-band and neuro-imaging information improved the specificity, but not the sensitivity or accuracy of the contact predictions [23]. Finally, T. Binder showed that the effect of DBS programming based on beta-frequency information did not significantly differ from effects achieved by image based programming when considering symptom control, TEED, programming time and stimulation amplitude [5].

The predictive accuracy of the second non-frequency factor, ERNA, was evaluated by three articles from research group #2. These studies reported predictive accuracies ranging from 68% to 80% [17, 20, 33].
