## Supplementary file 2 for "From subthalamic local field potentials to the selection of chronic deep brain stimulation contacts in Parkinson’s disease - A systematic review"

**Adapted JBI Critical Appraisal Checklist for Case Series [19]**
*(new or adjusted questions underlined)*

Reviewer……………………………………………………… Date…………………………………………..

Author………………………………………………………… Year…………………………………………..

| | Yes | No | Unclear | Not Applicable |
|---|---|---|---|---|
| 1. Were there clear criteria for inclusion in the case series? | ☐ | ☐ | ☐ | ☐ |
| 2. Was the LFP measurement performed in a standard, reliable way for all participants included? | ☐ | ☐ | ☐ | ☐ |
| 3. Were valid surgical methods described for all participants included in the case series? | ☐ | ☐ | ☐ | ☐ |
| 4. Did the case series have consecutive inclusion of participants? | ☐ | ☐ | ☐ | ☐ |
| 5. Was there clear reporting of the demographics of the participants in the study? | ☐ | ☐ | ☐ | ☐ |
| 6. Was there clear reporting of clinical information of the participants at the time of recordings (e.g. on/off)? | ☐ | ☐ | ☐ | ☐ |
| 7. Were the clinical outcomes and/or follow up results clearly reported? | ☐ | ☐ | ☐ | ☐ |
| 8. Was the physician blinded to LFP measurements during clinical contact evaluation in all patients? | ☐ | ☐ | ☐ | ☐ |
| 9. Was there clear reporting of the presenting site(s)/ clinic(s) demographic information? | ☐ | ☐ | ☐ | ☐ |
| 10. Was statistical analysis appropriate? | ☐ | ☐ | ☐ | ☐ |
| 11. Was there clear reporting of LFP acquisition/analysis? | | | | |
| a. LFP acquisition | | | | |
| i. Recording system and lead type reported? | ☐ | ☐ | ☐ | ☐ |
| ii. Sampling frequency reported? | ☐ | ☐ | ☐ | ☐ |
| iii. Participant state/position reported? | ☐ | ☐ | ☐ | ☐ |
| iv. Length of data segments reported? | ☐ | ☐ | ☐ | ☐ |
| v. Number of data segments reported? | ☐ | ☐ | ☐ | ☐ |
| b. Data analysis | | | | |
| i. Time-frequency conversion technique reported? | ☐ | ☐ | ☐ | ☐ |
| ii. Artefact extraction performance (y/n) reported? | ☐ | ☐ | ☐ | ☐ |
| iii. Normalisation performance (y/n) reported? | ☐ | ☐ | ☐ | ☐ |

Additional findings influencing study quality?

_______________________________________________________________________________

_______________________________________________________________________________

**Supplementary table 1.** Quality assessment sub-scores per included study according to the adapted JBI Critical Appraisal Checklist for Case Series.

| Article | JBI: Yes (Q1-3; Q4-8; Q9-10; Q11a-b) | JBI: No | JBI: Unclear |
|---|---|---|---|
| W. Dong (2023) [36] | 9 (2; 2; 1; 4) | 6 | 3 |
| J. Busch (2023) [34] | 15 (3; 4; 1; 7) | 2 | 1 |
| T. Binder (2023) [5] | 11 (3; 4; 1; 3) | 6 | 1 |
| J. Strelow (2023) [29] | 16 (3; 4; 1; 8) | 1 | 1 |
| B. Swinnen (2023) [28] | 9 (2; 3; 0; 4) | 6 | 1 (2 NA) |
| S. Lewis (2023) [27] | 15 (3; 4; 1; 7) | 2 | 1 |
| L. di Biase (2022) [23] | 12 (3; 2; 1; 6) | 4 | 2 |
| A. Shah (2023) [21] | 17 (3; 5; 1; 8) | 1 | 0 |
| J. Strelow (2022) [30] | 16 (3; 4; 1; 8) | 1 | 1 |
| S. Xu (2022-2) [20] | 12 (3; 4; 1; 4) | 6 | 0 |
| C. Chen (2022) [31] | 13 (3; 3; 1; 6) | 3 | 2 |
| C. Fernandez (2022) [35] | 15 (3; 4; 1; 7) | 2 | 1 |
| S. Xu (2022-1) [17] | 12 (2; 3; 1; 6) | 4 | 2 |
| N. Sinclair (2021) [33] | 13 (3; 3; 1; 6) | 3 | 2 |
| Y. Thenaisie (2021) [24] | 10 (3; 2; 0; 5) | 4 | 1 |
| I. Tamir (2020) [25] | 14 (3; 3; 1; 7) | 3 | 1 |
| T. Nguyen (2020) [15] | 11 (2; 2; 0; 7) | 5 | 2 |
| L. Milosevic (2020) [37] | 13 (2; 4; 1; 6) | 4 | 1 |
| R. Kochanski (2019) [26] | 15 (2; 4; 1; 8) | 2 | 1 |
| G. Tinkhauser (2017) [32] | 12 (1; 3; 1; 7) | 5 | 1 |
| A. Horn (2017) [22] | 14 (3; 3; 1; 7) | 3 | 1 |
| A. Connolly (2015) [16] | 11 (2; 3; 0; 6) | 5 | 2 |
| F. Yoshida (2010) [11] | 12 (2; 4; 1; 5) | 5 | 1 |
| N. Ince (2010) [1] | 14 (3; 4; 1; 6) | 3 | 1 |
| C. Chen (2006) [13] | 15 (3; 4; 1; 7) | 2 | 1 |
| **Median score (range)** | **13 (9-17)** | | |

*<u>Abbreviations</u>: JBI: Joanna Briggs Institute ; Q: question; NA: not applicable.*

**Supplementary table 2.** Extensive overview of relevant study characteristics and methodologies for the reviewed articles.

| **Article** | **City** (country) | **N STN** | **Recording** (system) | **Lead** (type) | **LFP measurements** | **Pre-processing** | **Feature(s)** (frequency range) | **Clinical reference** | **Prediction** (paradigm) |
|---|---|---|---|---|---|---|---|---|---|
| Dong (2023)[36] | Nanjing (China) | 72 | Externalised (MP-150 BIOPAC Systems, Inc) | Traditional (Type unknown) | Bipolar<br>- Intra-op. at rest<br>- Awake, OFF medication<br>- All rings | Artefact removal<br>- Notch filters<br>Normalisation<br>- Z-scoring | β/α-ratio z-score (12:30Hz/8:12Hz)<br>γ z-score (50:200Hz) | Chronic<br>- 6m. post-op.<br>- Cathode | Ranking (power max.) |
| Busch (2023)[34] | Berlin / Düsseldorf (Germany) | 32 | Internalised (Percept™ PC, Medtronic) | Directional (Sensight™, Medtronic) | Bipolar*<br>- 3m. post-op. at rest + during active contra-lateral hand movement<br>- Awake, OFF medication<br>- 2 non-adjacent ring and 3 ring crossing segments (BrainSense™ Survey) | Artefact removal<br>- High/low-pass filter<br>- Visual inspection | β-supp./peak (13:35Hz) | MPR<br>- 3m. post-op.<br>- Blinded to LFP<br>- Cathode | Ranking (peak supp. /power max.) |
| Binder (2023)[5] | Würzburg (Germany) | 16 | Internalised (Percept™ PC, Medtronic) | Directional (Sensight™, Medtronic) | Bipolar<br>- 3m. post-op. at rest<br>- Awake, OFF medication<br>- All segments (BrainSense™ Survey) | - | β-peak† (13:30Hz)<br>Low-β-peak (In case of more than one peak, 13:20Hz)<br>vs<br>Anatomy (image based) | MPR<br>- 3m. post-op.<br>- Blinded to LFP<br>- Cathode | Ranking (power max.) |
| Strelow (2023)[29] | Cologne / Jülich (Germany) | 13 | Internalised (Percept™ PC, Medtronic) | Directional (Sensight™, Medtronic) | Bipolar<br>- 3m. post-op. at rest<br>- Awake, OFF medication<br>- 2 non-adjacent rings (BrainSense™ Streaming) | Artefact removal<br>- band-pass filters<br>- Visual inspection<br>Normalisation<br>- Percentage of total power all frequencies | Low-β-supp. (13:20Hz)<br>High-β-supp. (21:35Hz) | MPR<br>- 3m. post-op.<br>- Blinded to LFP<br>- Cathode | Ranking (peak supp.) |
| Swinnen (2023)[28] | Amsterdam (Netherlands) | 8 | Internalised (Percept™ PC, Medtronic) | Traditional (3389, Medtronic) | Bipolar<br>- 4-7y. post-op. at rest (after replacement)<br>- Awake, ON medication<br>- All rings (BrainSense™ Survey) | - | β-peak (13:30Hz) | Chronic<br>- 4-7y. post-op.<br>- Cathode | Ranking (power diff.) |

| | | | | | | | | | |
|---|---|---|---|---|---|---|---|---|---|
| Lewis (2023)[27] | Aurora (CO, USA) Minneapolis (MN, USA) | 9 | Internalised (Percept™ PC, Medtronic) | Traditional (3389, Medtronic) | Bipolar<br>- ≥5ys. post-op. at rest (after replacement)<br>- Awake, OFF medication<br>- All rings (BrainSense™ Survey) | Normalisation<br>- rescaling from 0-1 | β-peak (13:30Hz) | Chronic<br>- ≥5ys. post-op.<br>- Blinded to LFP<br>- Cathode | Ranking (power diff.) |
| di Biase (2022)[23] | Rome / Pozzilli (Italy) Dubai Silicon Oasis (Dubai) | 28 | Externalised (SystemPLUS 98, Micromed S.p.A.) | Directional (Vercise Cartesia™, Boston Scientific) Traditional (n=1: Vercise, Boston Scientific) | Monopolar<br>- Intra-op. at rest<br>- Awake + under GA<br>- All rings and segments individually<br>- Cannula as reference | Artefact removal<br>- Notch & band-pass filters<br>Normalisation<br>- Divided by total power all frequencies | β-peak (13:29Hz) vs Anatomy (image based) | Chronic<br>- 1y. post-op.<br>- Cathode | Ranking (power max.) |
| Strelow (2022)[30] | Cologne (Germany) | 14 | Internalised (Percept™ PC, Medtronic) | Directional (Sensight™, Medtronic) | Bipolar<br>- 3m. post-op. at rest<br>- Awake, OFF medication<br>- All rings and segments (BrainSense™ Survey) | Artefact removal<br>- Visual inspection<br>- Perceive Toolbox<br>Normalisation<br>- Divided by total power across entire frequency range | β-peak (13:35Hz)<br>Low-β-av. (when no peak, 13:20Hz) | MPR<br>- 3m. post-op.<br>- Blinded to LFP<br>- Cathode | DETEC algorithm (power diff. + distance correction) |
| Chen (2022)[31] | Taoyuan (Taiwan) | 50 | Externalised (TMSi-Porti Amplifier, Twente Medical Systems International) | Traditional (3389, Medtronic) | Monopolar<br>- 5d. post-op. at rest<br>- Awake<br>- All rings individually<br>- common average as reference | Artefact removal<br>- High-pass + notch filters<br>Normalisation<br>- Dividing by total power all frequencies | θ-peak (4:7Hz)<br>α-peak (7:13Hz)<br>Low-β-peak (13:20Hz)<br>High-β-peak (20:35Hz)<br>Low-γ-peak (40:55Hz) | MPR<br>- 1m. post-op.<br>- Cathode | Ranking (power max.) |
| Xu (2022-2)[20] | (East) Melbourne / Heidelberg | 92 | Externalised (System unknown) | Traditional (3387, Medtronic) | Monopolar<br>- Intra-op. at rest<br>- Awake<br>- All rings individually | Normalisation<br>- Machine learning only: dividing by total power across | β-av. (13:30Hz)<br>ERNA | Chronic<br>- 6m. post-op.<br>- Blinded to LFP<br>- Cathode | Machine learning (Random forest) vs. |

| | | | | | | | | | |
|---|---|---|---|---|---|---|---|---|---|
| | / Fitzroy / Parkville (VIC, Australia) | | | | - Contra-lateral rings averaged as reference | contacts in hemisphere | (10x1/s: 3.375mA, 60µs, 130Hz) Anatomy (Eucl. distance) | | Ranking (power max.) |
| Fernandez-Garcia (2022)[35] | Madrid (Spain) | 28 | Externalised (Custom made) | Directional (Vercise Cartesia™, Boston Scientific) | Bipolar<br>- Intra-op. at rest<br>- Awake, OFF medication<br>- Between highest/lowest ring and every superior/inferior segmented electrode | Artefact removal<br>- Visual inspection<br>Normalisation<br>- Dividing by total power across contacts in hemisphere | Low-β-peak (13:20Hz)<br>High-β-peak (20:35Hz)<br>γ-peak (60:90Hz) | MPR<br>- 14-20d. post-op.<br>- Blinded to LFP<br>- Cathode | Ranking (power max.) |
| Shah (2022)[21] | Bern (Switzerland)<br>Oxford / Edinburgh (UK) | 27 | Externalised (TMSi-Porti Amplifier, Twente Medical Systems International) | Directional (Vercise Cartesia™, Boston Scientific) | Monopolar<br>- Intra-op. at rest + during active contra-lateral upper limb movement<br>- Awake, OFF medication<br>- All rings and segments individually<br>- common average as reference | Normalisation<br>- Z-scoring | α z-score (8:12Hz)<br>Low-β z-score (13:20Hz)<br>High-β z-score (21:30Hz)<br>γ z-score (60:90Hz)<br>Fast-γ z-score (105:145Hz)<br>HFO z-score (205:395 Hz)<br>Anatomy (Eucl. distance to sweet spot) | MPR<br>- 4-10m. post-op.<br>- Blinded to LFP<br>- Cathode | Feature selection + Ranking (Lasso regression + power max. of five best spectral features) |
| Xu (2022-1)[17] | (East) Melbourne / Heidelberg / Fitzroy / Parkville (VIC, Australia) | 28 | Externalised (g.USBamp, g.tec medical engineering GmbH) | Traditional (3387, Medtronic) | Monopolar<br>- Intra-op. at rest<br>- Awake<br>- All rings individually<br>- Contra-lateral rings averaged as reference | - | β-av. (13:30Hz)<br>HFO-av. (200:400Hz)<br>ERNA (10x1/s: 3.375mA, 60µs, 130Hz)<br>Anatomy | Chronic<br>- 3-18m. post-op.<br>- Blinded to LFP<br>- Cathode | Ranking (power max.) vs. Regression (Akin to 'best subset' regression) |

| | | | | | | | | | |
|---|---|---|---|---|---|---|---|---|---|
| | | | | | | | (Eucl. distance to sweet spot) | | |
| Then aisie (2021 )[24] | Lausanne (Switzerland) Würzburg (Germany) Leiden/ The Hague (Netherlands) | 19 | Internalised (Percept™ PC, Medtronic) | Traditiona l (3389, Medtronic) | Bipolar<br>- 2d-8m. post-op. at rest<br>- 5-8y. post-op. at rest (after replacement)<br>- Awake, OFF medication<br>- All rings (Survey) or all non-adjacent rings (Indefinite Streaming). | - | β-peak (13:35Hz) | MPR, n=17<br>- 2d-8m. post-op.<br>- Cathode<br><br>Chronic, n=2 (replacement)<br>- 5-8y. post-op.<br>- Cathode | Ranking (power max.) |
| Sincl air (2021 )[33] | (East) Melbourne/ Heidelberg / Parkville (VIC, Australia) | 24 | Externalised (g.USBamp, g.tec medical engineering GmbH) | Traditiona l (3387, Medtronic) | Monopolar<br>- Intra-op. at rest<br>- Awake + under GA<br>- All rings individually | Artefact removal<br>- Visual inspection | θ- + α-peak (4:12Hz)<br>β-peak (13:30Hz)<br>HFO-peak (200:400 Hz)<br>ERNA (10x1/s: 3.375mA, 60μs, 130Hz) | Chronic<br>- 1y. post-op.<br>- Blinded to LFP<br>- Cathode | Ranking (power max.) |
| Tamir (2020 )[25] | San Francisco (CA, USA) | 12 | Externalised (Microguide Pro™, Alpha Omega Engineering Ltd.) | Traditiona l (Vercise, Boston Scientific) | Monopolar‡<br>- Intra-op. at rest<br>- Awake, OFF medication<br>- Adjacent rings at various depths | Artefact removal<br>- Visual inspection<br>Normalisation<br>- Dividing by average baseline power (140-160Hz, no artefacts) | β-peak (13:30Hz) | Chronic<br>- 1y. post-op.<br>- Cathode | Ranking (power max.) |
| Nguy en (2020 )[15] | Bern/ Lausanne (Switzerland) | 4 | Externalised (LeadPoint system, Medtronic) | Direction al (directSTN ™ Acute, Aleva Neurothera peutics) | Monopolar<br>- Intra-op. at rest + during passive arm movement<br>- Awake, OFF medication<br>- 3 segments individually<br>- Cannula as reference | Artefact removal<br>- Visual inspection<br>Normalisation<br>- Dividing by average power across three segments. | Low-β-AUC (13:20Hz)<br>High-β-AUC (20:35Hz)<br>β-peak (13:35Hz) | MPR<br>- Intra-op.<br>- Cathode | Ranking‡ (power max.) |
| Milos evic | Tübingen (Germany) | 45 | Externalised (Neuro Omega™, | Traditiona l | Monopolar<br>- Intra-op. at rest<br>- Awake, OFF medication | Normalisation<br>- dividing by highest β-peak amplitude | β-peak (13:30Hz) | MPR<br>- ≥2m. post-op. | Ranking (power max.) |

| | | | | | | | | | |
|---|---|---|---|---|---|---|---|---|---|
| (2020)[37] | | | Alpha Omega Engineering Ltd.) | (3389, Medtronic (19); Infinity™, Abbott (26)) | - All rings individually at varying depths | peak (or average non-peak power for segments) recorded across all depths | | - Blinded to LFP<br>- Cathode and anode | |
| Kochanski (2019)[26] | Chicago (Ill, USA) | 10 | Externalised (Neuro Omega™, Alpha Omega Engineering Ltd.) | Traditional (3389, Medtronic (9); Infinity™, Abbott (1)) | Bipolar<br>- Intra-op. at rest + during active contra-lateral hand movement<br>- Awake, OFF medication | Artefact removal<br>- Visual inspection<br>Normalisation<br>- Substraction of 7th order polynomial fit to correct for the 1/f noise pattern | β-peak (13:30Hz) | MPR<br>- w(s)./m(s). post-op.<br>- Blinded to LFP<br>- Cathode | Ranking (power max.) |
| Tinkhauser (2017)[32] | Oxford (UK)<br>Bern/ Zürich (Switzerland)<br>Rome (Italy) | 19 | Externalised (ISIS IOM, inomed Medizintechnik GmbH) | Directional (Vercise Cartesia™, Boston Scientific) | Monopolar<br>- Intra-op. at rest<br>- Awake, OFF medication<br>- All segments individually<br>- Cannula as reference | Artefact removal<br>- Visual inspection<br>Normalisation<br>- Dividing by average power across entire β -band. | β-peak (13:30Hz)<br>Low-β-av. (when no peak, 13:20Hz) | MPR<br>- ≥4m. post-op.<br>- Blinded to LFP<br>- Cathode | Ranking (power max.) |
| Horn (2017)[22] | Berlin (Germany)<br>Boston (MA, USA) | 95 | Externalised (System unknown) | Traditional (3389, Medtronic) | Bipolar<br>- 2-7d. post-op. at rest<br>- Awake, OFF medication<br>- Adjacent rings | Artefact removal<br>- Visual inspection<br>- Notch filters<br>Normalisation<br>- Z-score | α z-score (7:13Hz)<br>Low-β z-score (12:20Hz)<br>High-β z-score (20:35Hz)<br>β z-score (13:35Hz) | Chronic<br>- 2-15 Ys. post-op.<br>- Cathode | Ranking (power max.) |
| Connolly (2015)[16] | Minneapolis (MN, USA) | 28 | Internalised (Activa® PC+S, Medtronic) | Traditional (3389, Medtronic) | Bipolar<br>- Up to 6m. post-op. at rest<br>- Awake, OFF medication<br>- All rings | Artefact removal<br>- Notch filters | θ-av. (3:5Hz)<br>α-av. (5:10Hz)<br>Low-β-av. (10:20Hz)<br>High-β-av. (20:30Hz)<br>γ-av. (60:90Hz) | Chronic<br>- ≤6m. post-op.<br>- Cathode | Machine learning + feature selection (Support vector machine + Leave-one-feature-out ) vs. |

| | | | | | | | | | |
|---|---|---|---|---|---|---|---|---|---|
| | | | | | | | | | Ranking<br>(power max.) |
| Yoshida (2010)[11] | London<br>(UK)<br>Fukuoka<br>(Japan)<br>Valencia<br>(Spain)<br>Taipei<br>(Taiwan) | 54 | Externalised<br>(System unknown) | Traditional<br>(3389, Medtronic) | Monopolar⸸<br>- Intra-op. at rest<br>- Awake, OFF medication<br>- Adjacent rings at various depths | - | β-peak<br>(11:35Hz) | Chronic<br>- ≥6m. post-op.<br>- Blinded to LFP<br>- Cathode | Ranking<br>(≥100% step change in max. power) |
| Ince (2010)[1] | Minneapolis<br>(MN, USA)<br>Atlanta<br>(GA, USA) | 4 | Externalised<br>(System unknown) | Traditional<br>(3389, Medtronic) | Monopolar⸸<br>- Intra-op. at rest + during active hand movement<br>- Awake, OFF medication<br>- Adjacent rings | Normalisation<br>- Dividing by total power across contacts | θ-peak<br>(3:7Hz)<br>α-peak<br>(7:13 Hz)<br>β-peak<br>(13:32Hz)<br>γ-peak<br>(48:220Hz) | MPR<br>- 3w. post-op.<br>- Blinded to LFP<br>- Cathode and anode | Ranking<br>(power max.) |
| Chen (2006)[13] | London<br>(UK)<br>Taipei<br>(Taiwan)<br>St. Petersburg<br>(Russia) | 16 | Externalised<br>(Biopotential Analyser Diana, IEPhB st. Petersburg) | Traditional<br>(3389, Medtronic) | Monopolar⸸<br>- Intra-op. at rest<br>- Awake, OFF medication<br>- Adjacent rings at various depths | Artefact removal<br>- Visual inspection | θ- + α-peak<br>(4:10Hz)<br>β-peak<br>(13:35Hz)<br>γ-peak<br>(65:85Hz) | MPR<br>- 6m. post-op.<br>- Blinded to LFP<br>- Cathode | Ranking of best two<br>(power max.) |

*: reconstruction of monopolar segments from bipolar (1 non-adjacent ring and 3 contact-level crossing segments) recordings using custom formula; † : beta-suppression also investigated but only compared to beta-peak and not to MPR or image-based programming; ‡ : the three beta-features were additionally combined in a simple ranking method which provided higher performance scores than the individual beta-features alone; ⸸ : online bipolar re-referencing of monopolar recordings (supplementary results, Supplementary file 1). *Abbreviations N STN: number of STN; LFP: local field potential; intra-op.: intra-operative; post-op.: post-operative; d.: days; w.: weeks; m.: months; y.: years; HFO: high frequency oscillations; ERNA: evoked resonant neural activity; max.: maximum; diff.: difference; GA: general anesthesia; LASSO: least absolute shrinkage and selection operator.*

**Supplementary table 3**. Prediction of the two most optimal contacts based on beta-frequency oscillations alone or in combination with other (non-) frequency factors compared to the clinical stimulation contact.

| Article | Prediction methods | Outcome | Number of contacts | Score | Number of STN | Medica-tion state | Clinician blinding | JBI score (Y/N/U) |
|---|---|---|---|---|---|---|---|---|
| One beta-oscillation factor alone | | | | | | | | |
| Monopolar recordings | | | | | | | | |
| [21]‡ | Predictive accuracy low-β-peak | 56.0% (TW) | 8 | Moderate | 27 | OFF | Y | 17/1/0 |
| [32] | Predictive accuracy β-peak | 92.0% (CE) | 6 | Moderate | 19 | OFF | Y | 12/5/1 |
| [37] | Agreement β-peak | 93.3% | 4# | Low | 45 | OFF | Y | 13/4/1 |
| [17] | Agreement average-β-power | 89.0% | 4 | Low | 28 | U | Y | 12/4/2 |
| [21]‡ | Predictive accuracy low-β-peak | 64.0% (TW) | 6 | Low | 27 | OFF | Y | 17/1/0 |
| Pseudomonopolar recordings *(bipolar LFP recordings transformed to monopolar configuration using custom technique)* | | | | | | | | |
| [34]‡ | Predictive accuracy β-suppression | 75.0% | 6 | Moderate | 32 | OFF | Y | 15/2/1 |
| [34]‡ | Predictive accuracy β-peak | 50.0% | 6 | Low | 32 | OFF | Y | 15/2/1 |
| [30]‡ | Predictive accuracy β-peak | 78.6% (CE) | 3 | Low | 14 | OFF | Y | 16/1/1 |
| [30]‡ | Predictive accuracy β-peak | 71.4% (CE) | 4 | Low | 14 | OFF | Y | 16/1/1 |
| Bipolar recordings | | | | | | | | |
| [35] | Correlation β-peak | r=-0.68, p=0.0001 | 8 | Moderate | 28 | OFF | Y | 15/2/1 |
| [29]‡ | Predictive accuracy low-β-suppression | 67.0% | 8 | Moderate | 13 | OFF | Y | 16/1/1 |
| [29]‡ | Predictive accuracy high-β-suppression | 50.0% | 8 | Low | 13 | OFF | Y | 16/1/1 |
| [13] | Agreement β-peak | 83.0% | 4 | Low | 16 | OFF | Y | 15/2/1 |
| Machine learning/multi-factor using multiple frequency bands | | | | | | | | |
| Monopolar recordings | | | | | | | | |
| [21]‡ | LASSO (α,low/high-β,(fast)-γ,HFO) | 58.0% (TW) | 6 | Low | 27 | OFF | Y | 17/1/0 |
| [21]‡ | LASSO (α,low/high-β,(fast)-γ,HFO) | 47.0% (TW) | 8 | Low | 27 | OFF | Y | 17/1/0 |

Performance score: High: >3 times a priori chance; moderate: ≤3 times but >2 times a priori chance OR significant correlation with p<0.005; low: ≤2 times but > 1 time a priori chance OR significant correlation with p<0.05; poor: ≤ 1 time a priori chance OR non-significant correlation with p>0.05; #: both contacts from bipolar stimulation configuration used as reference for predictions (supplementary results, Supplementary file 1); ‡: multiple outcomes presented in the table.
*<u>Abbreviations:</u> Y: Yes; N: No; U: Unknown/Unclear; STN: subthalamic nucleus; HFO: High frequency oscillation; TW: contact with largest therapeutic window (difference in stimulation current between effect threshold and side-effect threshold) used as reference for prediction; CE: contact with highest clinical efficiency*

*(stimulation induced improvement in contralateral MDS-UPRS-III (sub)scores OFF-medication) used as reference for prediction; MDS-UPDRS-III: Movement Disorder Society Unified Parkinson's Disease Rating Scale part three; HFO: High frequency oscillation; LASSO: Least Absolute Shrinkage and Selection Operator regression.*
